# The path from brain to words is rerouted in opioid addiction

**DOI:** 10.64898/2026.10.24.26366318

**Authors:** Shani Pais, Refael Tikochinski, Nitay Calderon, Sarah G. King, Rita Z. Goldstein, Roi Reichart

## Abstract

Whether spontaneous speech preserves how the brain encoded an experience is unknown. We tested this question in opioid use disorder (OUD), in which drug cues outweigh other rewards. Eighty-two people with OUD and 34 controls watched the opening of the film Trainspotting during functional magnetic resonance imaging, then retold it in their own words. Language-model analyses showed that people with OUD dwelt on craving, personal identification and recovery, devoting more of their retelling to drug scenes. In 65 participants with imaging, this speech bias tracked how strongly the brain had distinguished drug from non-drug scenes during viewing, in left frontoparietal cortex in controls but left temporal cortex in OUD. Direct comparison showed left accumbens coupling positive in OUD and negative in controls, and left superior frontal coupling weaker in OUD. Spontaneous speech thus carries a trace of neural encoding whose route depends on what the experience meant to the speaker.

## 1 Introduction

What a person says about an experience is shaped by how their brain encoded it. Naturalistic neuroimaging has shown that spoken recall during scanning can reinstate the neural patterns established at encoding [1–3] but the link between encoding and the spontaneous description that follows, outside the scanner, is rarely measured directly. Unconstrained speech unfolds over extended timescales, integrates perception with memory and emotion, and engages distributed neural systems supporting self-referential, social, and executive processes [4, 5]. Constrained laboratory paradigms have defined much of what is known about encoding and retrieval, yet their structure limits generalizability and clinical utility. Whether what a person says about an experience, in their own words and within the hour, preserves how their brain encoded it is unknown. If it does, speech becomes a readout of encoding, and people for whom the same content carries different meaning become a test of whether that readout runs through the same systems.

Drug addiction is the decisive test case: the same film carries different meaning for people with and without the disorder. Addiction models predict that drug content captures salience at the expense of other content and is weighted as self-relevant [6–8], so if speech preserves encoding, the route from one to the other should differ by group. In the impaired Response Inhibition and Salience Attribution (iRISA) framework, this salience bias is coupled with diminished engagement of control and self-regulatory systems, reflected in exaggerated drug-cue responses across reward, salience, default mode, and control networks [9]. Its core tenet is a seesaw: because the value of a cue depends on what it competes with [10], drug cues outcompete other arousing reinforcers (such as food and sexual cues) sharing the same context, calling for designs that contrast drug content against non-drug content embedded in the same narrative rather than against neutral cues [11]. These neural biases track craving and relapse risk [12, 13].

Recent work has turned to complex, real-world stimuli, such as narrative films, during functional MRI [14]. Such paradigms elicit reliable, drug-biased neural responses in individuals with substance use disorders, and those responses can change with treatment. In everyday life, people do not passively view drug cues in isolation; they remember, recount, and reinterpret them through language. Spontaneous speech carries addiction-relevant signals that predict subsequent drug use [15]. In the cohort studied here, the similarity structure of recall across participants tracks the similarity structure of neural responses during viewing [16], an analysis defined over whole transcripts and whole time series. Whether spontaneous speech carries a behavioral trace of the prior neural encoding of drug content specifically, rather than of the narrative as a whole, remains unknown; if it does, a few minutes of speech would index the neural salience bias that tracks craving and relapse risk [12, 13].

Here we combine transformer-based language models with fMRI to test whether what people say about a drug-themed film, within 45 minutes of leaving the scanner, preserves how strongly their brains separated drug from non-drug scenes while they watched, and whether the same regions carry that link in people with opioid use disorder (OUD) and in healthy controls (HC). Both groups viewed the same addiction-themed movie clip during fMRI and afterwards recalled the narrative and their emotional responses to it in their own words. Recall was analyzed with two complementary approaches: data-driven topic modeling to characterize semantic content, and sentence-to-scene similarity to align speech with specific moments in the movie. We then asked whether individual differences in the relative emphasis on drug versus non-drug content in speech were coupled to the neural contrast between drug and non-drug scenes during viewing. Following iRISA, we predicted group differences in coupling along both of its axes, mesolimbic salience attribution and prefrontal control.

People with OUD retold the film differently from HC participants: they dwelt on craving, identification with the film and recovery, and gave drug scenes a larger share of their recall. This behavioral bias was coupled to neural activity recorded earlier, during viewing, and the coupling dissociated by group along the two axes specified by iRISA: positive in the left nucleus accumbens in OUD and negative in HC, strong in the left superior frontal gyrus in HC and reduced in OUD.

## 2 Results

### 2.1 Free recall speech reveals markers of opioid use disorder

A total of 82 participants with OUD (65 males, mean age = 41.5, SD = 10.2) and 34 HC participants (22 males, mean age = 40.8, SD = 11.0; see Table 1) viewed the same movie clip featuring heavy drug-use cues during functional MRI scanning (the first 17 minutes and 3 seconds of Trainspotting; see Methods). After the scan, participants were asked to recall the movie’s narrative and describe their experience of watching it, while their speech was recorded.

**Table 1:** Baseline demographic, neuropsychological, and drug use/addiction measures of participants in topic analysis.

|  | HC<br>N = 34 | Opioid use<br>disorder<br>OUD, N = 82 | Test<br>statistic | *p* value |
| --- | --- | --- | --- | --- |
| **Demographics** |  |  |  |  |
| Age, mean (SD) years | 40.8 (11.0) | 41.5 (10.2) | $U = 1322$ | .664 |
| Gender, # men/women | 22/12 | 65/17 | $\chi^2 = 2.00$ | .158 |
| Race, # Black/White/multiracial or other | 11/15/8 | 11/54/17 | $\chi^2 = 6.54$ | .038 |
| Education, mean (SD) years | 16.1 (2.9) | 11.9 (2.4) | $U = 2378$ | <.001 |
| **Neuropsychological tests** |  |  |  |  |
| WRAT - mean (SD) Reading standard score | 108.7 (7.2) | 96.4 (11.9) | $U = 2245$ | <.001 |
| WASI - mean (SD) Matrix Reasoning scaled score | 11.5 (2.9) | 10.2 (2.9) | $U = 1784$ | .018 |
| Depression symptoms, mean (SD) BDI score | 4.3 (9.3) | 12.6 (10.4) | $U = 502$ | <.001 |
| Anxiety symptoms, mean (SD) BAI score | 3.1 (6.4) | 9.9 (9.8) | $U = 617$ | <.001 |
| **Drug use and addiction variables** |  |  |  |  |
| Regular nicotine use, # current/past/never | 1/5/28 | 76/5/1 | – | – |
| Alcohol dependence, mean (SD) SMAST score | 0.3 (0.9) | 2.7 (3.1) | $U = 436$ | <.001 |
| *Heroin use disorder* |  |  |  |  |
| Age of onset, mean (SD) years | – | 24.5 (7.8) | – | – |
| Regular use, mean (SD) years | – | 11.9 (7.6) | – | – |
| Current abstinence, mean (SD) days | – | 201.1 (224.4) | – | – |
| Past 30-day use, mean (SD) days | – | 0.5 (3.3) | – | – |
| Craving symptoms, mean (SD) HCQ score | – | 40.2 (14.6) | – | – |
| Withdrawal symptoms, mean (SD) SOWS score | – | 3.3 (4.7) | – | – |
| Severity of dependence, mean (SD) SDS score | – | 11.3 (3.7) | – | – |
| Craving before the scan, mean (SD) 0–9 rating | – | 2.5 (2.2) | – | – |
| Scene-induced craving, mean (SD) maximum 0–9 rating | – | 5.4 (3.2) | – | – |

**Notes:** The table presents mean and standard deviation (SD) or quantities for baseline sample characteristics. Test statistics were derived from a Mann-Whitney U test ( $U$ ) for continuous variables and chi-squared test ( $\chi^2$ ) for categorical variables. Since only one HC participant reported current nicotine use and only 6 OUD participants reported non-current nicotine use, no statistical test was performed on this measure. Missing data (HC/OUD): education 1/1; BDI 0/2; BAI 0/2; alcohol dependence 0/1; regular heroin use 0/3; HCQ 0/1; SOWS 0/1; craving before the scan 0/2. Craving before the scan was rated in the scanner; scene-induced craving is the maximum of the fixed-schedule movie ratings (see Methods). The remaining measures reported in Supplementary Table S8 are craving after the scan (3.0 (2.5); $n = 80$ ), the pre-to-post change across the scan (0.5 (1.7); $n = 80$ ), the mean of the scene-induced ratings (1.1 (1.1); $n = 81$ ), and the session-to-session change in that mean (–0.4 (0.8); $n = 40$ ), which is derived from the second session and so is not a baseline measure.

#### 2.1.1 Topic Modeling

Using data-driven topic modeling, which groups sentences from the recall speech into coherent themes without predefined categories (BERTopic; see Methods), we identified 21 interpretable topics, each representing a cluster of semantically related sentences capturing a recurring theme (Supplementary Table S1). For each participant, we then quantified the prevalence of each topic.

Comparisons revealed significant differences in topic prevalence between groups (Table 2; see Methods). Four topics were significantly more prevalent among OUD than HC participants: “Personal feeling and identification with the movie” (Topic 2, *g* = 0.47, 95% *CI*_*BCa*_ [0.22, 0.67], *p*_corr_ = .015), “Triggered ‘getting high’ experiences and cravings” (Topic 7, *g* = 0.45, 95% *CI*_*BCa*_ [0.27, 0.58], *p*_corr_ = .015), “Personal usage craving and memories triggered by the movie” (Topic 11, *g* = 0.38, 95% *CI*_*BCa*_ [0.25, 0.49], *p*_corr_ = .008), and “Desire for rehabilitation” (Topic 13, *g* = 0.50, 95% *CI*_*BCa*_ [0.30, 0.66], *p*_corr_ = .015). No topic was significantly more prevalent among HC participants after correction.

**Table 2:**
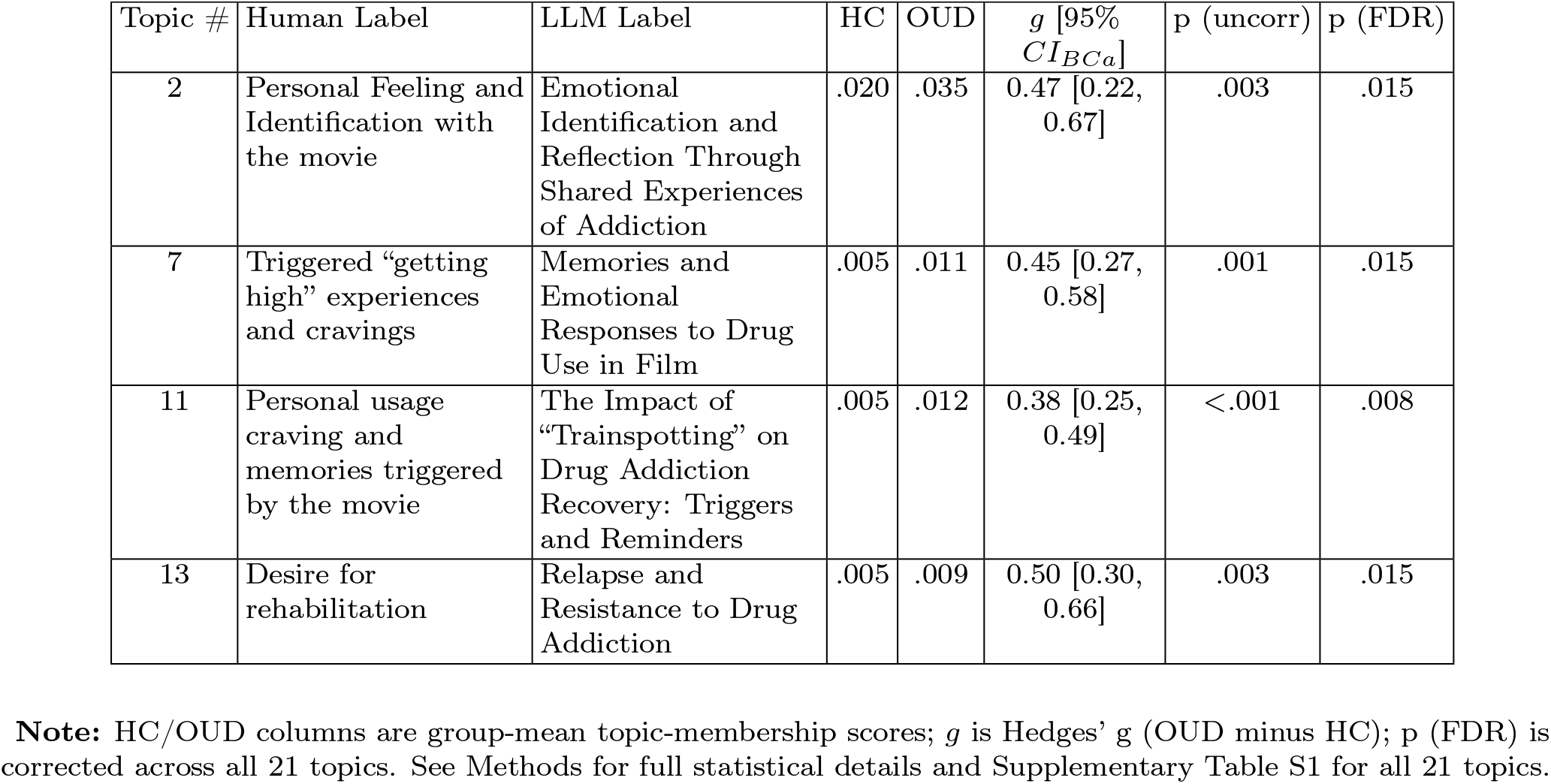
Significant Topics: permutation-test results and topic labeling.

### 2.1.2 Scene similarity

As a complementary, top-down analysis, we quantified how much of each participant’s recall was devoted to each of the 15 pre-defined movie scenes using sentence-to-scene similarity mapping (see Methods). In brief, each recall sentence was assigned to the scene whose textual description it most closely matched, and we then computed, for each participant, the proportion of sentences mapped to each scene.

At the individual scene level, two scenes showed higher average recall proportions among OUD than HC participants in an uncorrected comparison: Scene 4 (“Mother Superior”; *g* = 0.43, 95% *CI*_*BCa*_ [0.20, 0.62], *p* = .006) and Scene 8 (“Parent with drug addiction”; *g* = 0.35, 95% *CI*_*BCa*_ [−0.03, 0.60], *p* = .048). Both scenes were explicitly related to drug use, but neither survived correction across the 15-scene family.

To test the expected drug-bias effect directly, we used the a priori classification of each of the 15 scenes as drug-related (*n* = 7) or non–drug-related (*n* = 8) based on explicit drug use or drug presence (see Methods). This classification was previously applied only to the movie scenes themselves [14, 16]; here we applied it to speech for the first time, recalculating, for each participant, the proportion of sentences related to any of the drug-related scenes. This single, pre-specified contrast revealed a group difference: OUD participants devoted a larger proportion of their recall to drug-related scenes than the HC group (OUD: *M* = 0.634, *SD* = 0.19; HC: *M* = 0.501, *SD* = 0.205; *g* = 0.68, 95% *CI*_*BCa*_ [0.28, 1.10], *p* = .002) (Fig. 1). Within groups, OUD participants recalled drug-related scenes disproportionately relative to non–drug-related scenes (paired *t*(81) = 6.38, *d*_*z*_ = 0.71, *p <* .001), whereas the HC group allocated roughly equal attention to the two categories (paired *t*(33) = 0.04, *d*_*z*_ = 0.01, *p* = .97). The two proportions sum to one within each participant, so the group difference in the drug-scene proportion (*p* = .002) is equivalent to the group × scene-category interaction. Together with the topic-modeling findings, these results indicate that OUD participants were biased toward drug-related content and toward scenes personally and emotionally relevant to their own drug use. None of the group differences reported in this section was explained by the demographic, cognitive, or clinical variables that differed between groups (Table 1); adjusting for these variables left each effect intact (see Methods and Supplementary Results).

**Figure 1:**
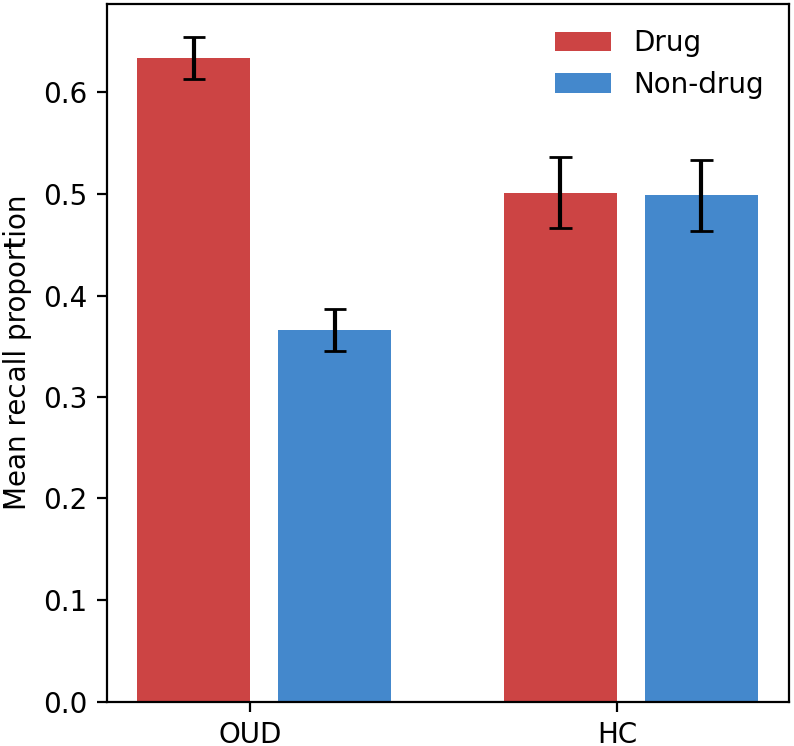
Mean proportion of recall sentences assigned to drug and non-drug scenes by group (HC vs. OUD), Session 1. Error bars indicate Standard Error of the Mean (SEM).

### 2.2 Drug-biased speech couples to accumbens encoding in OUD and to prefrontal encoding in HC

Given the drug-biased recall in the OUD group, we next asked whether this behavioral signature corresponded to brain activity during movie viewing. Neuroimaging analysis was restricted to participants with usable fMRI data who completed the entire study data collection, yielding 35 OUD and 30 HC participants (Supplementary Table S2), the same fMRI cohort analyzed in [16] and drawn from the same study as [14]. The drug-biased speech pattern identified in the full sample held in this subset: OUD participants devoted a larger proportion of recall sentences to drug scenes than HC participants (OUD: *M* = 0.618, *SD* = 0.17; HC: *M* = 0.492, *SD* = 0.22; *g* = 0.65, 95% *CI*_*BCa*_ [0.08, 1.08], *p* = .012).

We first analyzed neural activity at the level of the region of interest (ROI) using the averaged blood-oxygen-level-dependent (BOLD) signal of cortical areas [17]. For each ROI, we computed a participant-specific *Neural Contrast* : BOLD activation during drug scenes minus activation during non-drug scenes (see Methods for the full scene-wise processing). This measure reflects the degree to which each brain region responded preferentially to drug-related content during movie viewing. To link these neural patterns to behavior, we correlated the Neural Contrast across participants within each group with the language-AI-derived *Speech Contrast*, which quantifies the relative proportion of speech devoted to drug versus non-drug scenes during recall.

In the OUD group, coupling between the Speech Contrast (post-scanning recall) and the Neural Contrast (in-scanner viewing) was significant after network-wise FDR correction in three ROIs, all positive, all in the left hemisphere and anatomically clustered in the temporal cortex: the temporal pole (LH_Limbic_TempPole_7; *r* = 0.46, *p*_corr_ = .042) and two adjacent regions spanning the temporal pole and superior temporal gyrus (LH_TempPar_1; *r* = 0.41, *p*_corr_ = .035; LH_TempPar_2; *r* = 0.44, *p*_corr_ = .035). These effects point to left superior temporal cortex as the primary site of brain–speech coupling in OUD participants (full results, including uncorrected associations, in Supplementary Table S3).

In the HC group, coupling was significant after correction in 14 ROIs, all in the left hemisphere, spanning Control B (6 ROIs), Control A (4 ROIs), Default Mode A (2 ROIs), and Default Mode B (2 ROIs) networks. Correlations reached *r* = 0.68 in the left superior parietal lobule / intraparietal sulcus (LH_ContA_IPS_5; *p*_corr_ = .001), *r* = 0.62 in the left rostral superior frontal gyrus (LH_DefaultA_PFCd_2; *p*_corr_ = .002), and *r* = 0.62 in the left frontal pole (LH_DefaultB_PFCd_3; *p*_corr_ = .004). The full list of these ROIs is given in Supplementary Table S4 (uncorrected associations in Supplementary Table S5). Cortical surface maps of the FDR-corrected brain–speech coupling patterns are shown in Fig. 2a (OUD) and Fig. 2b (HC), and group differences in Fig. 2c. The within-group test was two-tailed, so negative correlations were equally testable; they were present at the uncorrected level in both groups but none survived FDR correction (Supplementary Tables S3 and S5). The two within-group ROI sets did not overlap: no region was significant in both groups.

**Figure 2:**
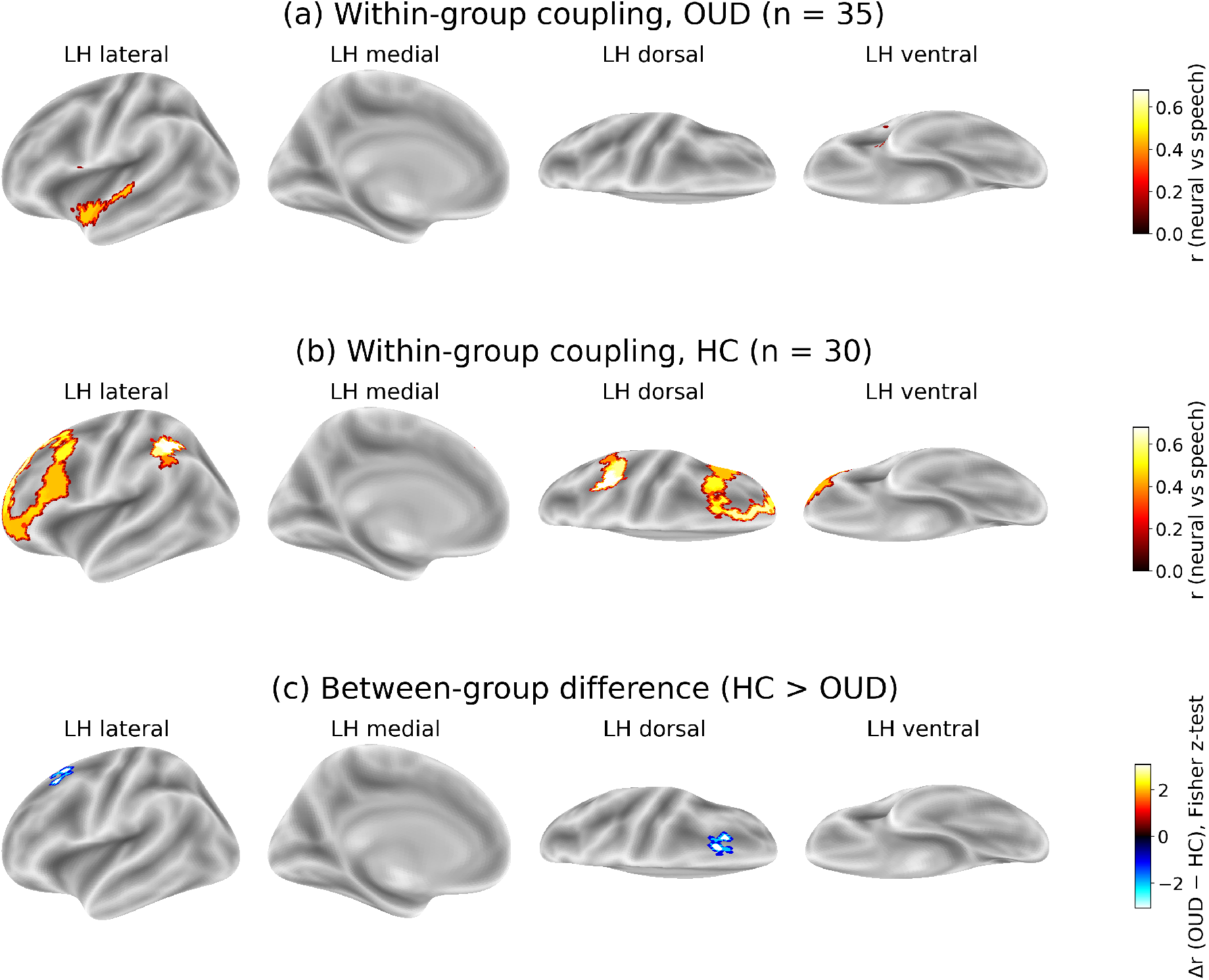
Cortical surface maps of brain–speech coupling (drug vs. non-drug contrast). **(a)** Regions in which the correlation between the Speech Contrast and Neural Contrast was significant after network-wise FDR correction in the OUD group; participants who devoted more recall speech to drug scenes showed greater neural differentiation of drug scenes in these regions. **(b)** The corresponding regions in the HC group. Panels (a) and (b) share a colour scale (*r*, neural vs. speech) so the two maps can be compared directly. **(c)** The single region with a significant group difference in coupling surviving network-wise FDR correction: left superior frontal gyrus within the Default Mode A network (LH_DefaultA_PFCd_3; *z* = −3.07, *p*_corr_ = .022), reflecting stronger coupling in HC; colour denotes the signed Fisher *z* difference on its own scale. Every region surviving correction in all three maps was left-hemisphere, and no region survived correction in the right hemisphere in either group or in the group comparison; right-hemisphere surfaces are therefore not shown. Full per-ROI results, including associations significant only at the uncorrected level, are listed in Supplementary Tables S3 (OUD), S4 and S5 (HC), and S6 (group differences).

For each ROI we then compared the Speech Contrast–Neural Contrast coupling between groups (Fisher r-to-z; see Methods). One cortical region showed a significant group difference after correction, with stronger coupling in the HC group: the left superior frontal gyrus within the Default Mode A network (LH_DefaultA_PFCd_3; *z* = −3.07, *p*_corr_ = .022). Within groups, this cortical coupling was strong and positive in HC (*r* = 0.55) and weakly negative in OUD (*r* = −0.19). The difference was not an artifact of one group varying less than the other: the spread of Speech and Neural Contrast values was comparable across groups in this region, and the difference held when the groups were matched on range (Supplementary Results). Full cortical results, including uncorrected group differences, are in Supplementary Table S6.

We repeated the coupling analysis over the 15 subcortical structures of the Harvard– Oxford atlas [18–21]. One structure showed a significant group difference in coupling after correction: the left nucleus accumbens (*z* = 3.55, *p*_corr_ = .001), the largest effect observed anywhere in the analysis, cortical or subcortical. The difference reflected oppositely signed within-group correlations, positive in OUD (*r* = 0.34) and negative in HC (*r* = −0.52; Fig. 3): OUD participants who recalled more drug scenes showed greater nucleus accumbens differentiation of drug scenes, whereas HC participants who did so showed less. Neither within-group correlation survived correction on its own (OUD *p*_corr_ = .361; HC *p*_corr_ = .054). Within-group correlations for all 15 structures are provided in Supplementary Table S7. Both corrected group differences survived adjustment for the variables that differed between groups and re-testing under range-matching (Supplementary Results), and the two run in opposite directions across cortex and striatum. We tested that reversal directly as a region × group interaction (see Methods): the difference between the accumbens and superior frontal gyrus group differences in coupling exceeded every one of 10,000 group-label permutations (*D* = 6.62, bootstrap 95% *CI* [4.09, 9.47], *p* = .0001), and a Neural Contrast × Group × Region term in a participant-clustered regression confirmed it (*β* = 1.75, *z* = 3.67, *p <* .001).

**Figure 3:**
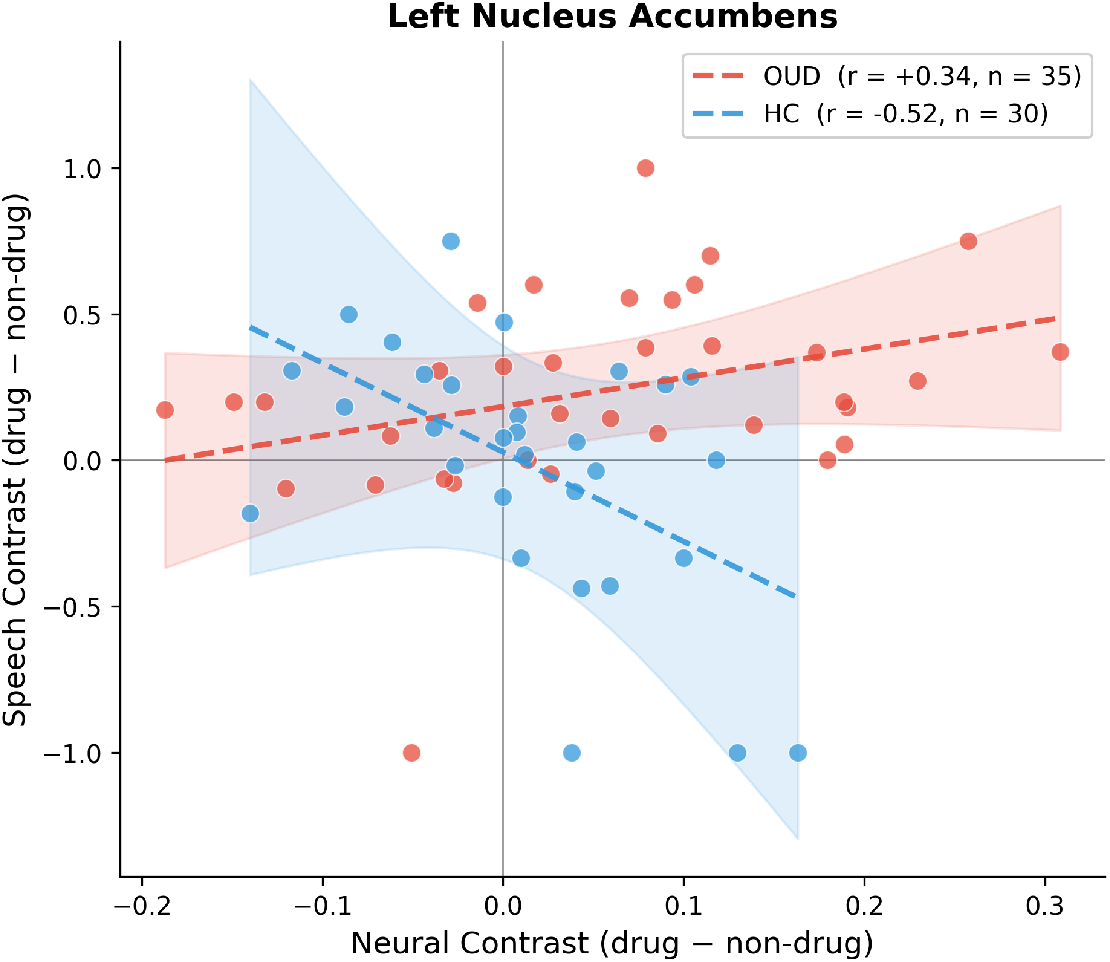
Speech Contrast–Neural Contrast coupling in the left nucleus accumbens, by group. Each point is one participant; the Neural Contrast (drug minus non-drug) is plotted against the Speech Contrast (drug minus non-drug), with dashed within-group regression lines and shaded 95% confidence bands. Coupling is positive in OUD (*r* = 0.34, *n* = 35) and negative in HC (*r* = −0.52, *n* = 30), the oppositely signed within-group correlations underlying the group difference (group *z* = 3.55, *p*_corr_ = .001; Supplementary Table S7).

We then related the five regions carrying the main results (left nucleus accumbens, left superior frontal gyrus, and the three left temporal regions coupled in OUD) to the 13 drug-use, craving, and withdrawal measures available in OUD, correcting across regions within each measure (see Methods). Craving reported before the scan (*n* = 34) interacted with the Speech Contrast in predicting the Neural Contrast in the temporal pole (LH_Limbic_TempPole_7; *β* = 0.30, *q* = .045) and the adjacent temporoparietal region (LH_TempPar_2; *β* = 0.40, *q* = .045). Neither effect survived refitting the moderation with the Speech Contrast as the outcome or a median-split comparison of the within-group correlations; we therefore do not interpret them as craving effects on coupling (Supplementary Results). Craving induced by the movie scenes (*n* = 35) did not moderate coupling anywhere, including the nucleus accumbens, where the interaction was flat and nominally opposite in sign, and the superior frontal gyrus (*p* = .70).

No other drug use measure survived correction against accumbens or prefrontal coupling. The nearest trends, accumbens coupling in participants with earlier onset of use (*q* = .053) and control-like prefrontal coupling with longer abstinence (*q* = .107), run in the expected directions but are too weak to interpret. Within the imaging subsample, the Speech Contrast was higher in participants with later onset of regular use (*ρ* = 0.38, *p* = .027, *n* = 35) and lower in those with longer duration of use (*ρ* = −0.38, *p* = .028, *n* = 34); neither association held in the full sample, so both are treated as subsample-specific. Results for all 13 measures, with the sample size for each, are given in Supplementary Table S8.

Scene-induced craving was sampled on a fixed 30-s schedule rather than at the drug scenes themselves, so we recomputed it a priori over only the clips falling inside the drug scenes used for the two contrasts (19 of 35 clips), together with dispersion variants and a within-subject drug-minus-non-drug difference (six variables, uncorrected; see Methods). In the full sample (*n* = 74 with per-clip ratings), the drug-clip maximum (*ρ* = 0.290, *p* = .011) and the within-subject drug-minus-non-drug difference (*ρ* = 0.266, *p* = .023) tracked the Speech Contrast; in the imaging subsample (*n* = 32), none of the six variables was associated with the Neural Contrast or moderated coupling in any region, the nucleus accumbens included (strongest *r* = 0.265, *p* = .136). The restriction did not change the measure (drug-clip vs. original mean, *r* = 0.961), so the fixed sampling schedule had not diluted drug-scene craving (Supplementary Table S9).

## 3 Discussion

What people said about the film, within 45 minutes of leaving the scanner, preserved how strongly their brains had separated drug from non-drug scenes while they watched it. In HC the link ran through left frontoparietal cortex and in OUD through left temporal cortex, and the groups diverged in opposite directions in the accumbens and the superior frontal gyrus: the salience and control limbs of iRISA. The two limbs are ordinarily inferred from separate tasks, often in separate samples; here both emerged from one measure in the same people.

Two complementary analyses converged: bottom-up topic modeling showed that participants with OUD over-represented themes of personal identification with drug content, triggered craving, and recovery, and top-down scene-similarity mapping showed disproportionate allocation of their verbal recall to drug-related scenes relative to HC. Both express the enhanced salience attribution to drug cues that models of addiction postulate, now measurable in what people say spontaneously.

Two group differences survived correction, and together they express the two limbs of the iRISA model in opposite directions within a single paradigm. In the left nucleus accumbens, a core substrate of salience attribution, coupling was positive in OUD and negative in HC, the largest effect in the study: the more a participant’s accumbens differentiated drug scenes during viewing, the more that participant’s later speech dwelt on them. In the left superior frontal gyrus, a dorsal prefrontal region, coupling was strong in HC and reduced in OUD: controls recruited prefrontal cortex to carry scene-specific drug encoding into later speech, and this prefrontal route was not evident in OUD, consistent with the diminished executive engagement highlighted as core to addiction by the iRISA model [6, 7, 9, 11]. That the striatal half appears in an opioid cohort fits evidence that ventral striatal drug-cue reactivity [9, 11] and grey matter volume reductions [22] are especially characteristic of OUD. Neither within-group accumbens correlation survived correction on its own (the HC superior frontal correlation did), so this reading rests on the two group differences themselves, both of which survived correction, covariate adjustment, and range-matching.

The two within-group maps point to different routes from viewing to speech. In OUD, participants who recalled more drug-related content showed greater neural differentiation of drug versus non-drug scenes in the left superior temporal cortex, including the temporal pole and superior temporal gyrus, regions supporting auditory-linguistic processing and the personal-semantic knowledge that underlies autobiographical retrieval [23–25]. In HC, the same brain-speech relationship engaged more regions (14 versus 3), spanning left frontoparietal cortex across Default Mode and Control networks implicated in narrative comprehension, scene segmentation, and memory consolidation [1–3].

The two routes are the two halves of iRISA. In HC, speech about drug scenes tracked the frontoparietal regions that track any scene, the externally directed route from viewing to speech. In OUD, speech tracked temporal regions tied to personal semantic memory, and in the accumbens the link ran opposite to HC: what participants later said reproduced a personally weighted encoding, not a weaker version of the HC pattern.

The shared film is what makes the speech comparable across people and the drug bias interpretable: drug and non-drug content compete within one story, under one instruction, so what a participant chose to say is an outcome of that competition rather than a reaction to a cue in isolation. It also removes the variability that ordinarily confounds naturalistic speech paradigms, where topic and eliciting conditions differ from person to person, while retaining the ecological validity of unconstrained recall. Movie-elicited recall is in this sense a “standardized free speech” paradigm, analogous to the picture-description tasks used in aphasia, dementia, and Alzheimer’s disease research [26– 28], but with a dynamic, emotionally complex and disorder-relevant stimulus in place of fixed, mostly neutral pictures. The same logic has precedent in health: readers assigned the perspective of a burglar or a home buyer later recalled the details relevant to that perspective [29]; here the perspective is supplied by the disorder.

Methodologically, transformer-based language models can extract clinically meaningful semantic structure from brief, unconstrained speech samples. Prior applications in psychiatry have relied on clinical notes and structured interview responses [30], social media posts [31, 32], or naturalistic speech (about consequences of use) in addiction [15]. Such sources capture naturalistic expression but lack a shared stimulus or a controlled environment, or a verified diagnostic status in deeply phenotyped individuals. Semantic embeddings and topics derived from a few minutes of narrative recall, collected with minimal instruction, reveal systematic group differences that align with theoretical predictions and correspond to the underlying neural process.

The neuroimaging contribution builds on two prior analyses of this cohort. Kronberg et al. [14] reported group-level drug-biased neural synchronization to this movie in this cohort, localized to orbitofrontal cortex and recovering with treatment; that analysis was defined over the group, did not detect subcortical results, consistent with other naturalistic studies that have generally not analyzed subcortical areas. King et al. [16] related whole-transcript semantics to whole-movie neural similarity across pairs of participants in the same cohort, a second-order measure blind by construction to which stimulus content drives the correspondence. Neither analysis relates drug-specific speech to drug-specific neural response in the same person. The coupling reported here is first-order: one drug versus non-drug contrast per participant on both the brain and behavior sides, correlated across participants. Linking neural substrates to speech in this way addresses a persistent translational gap in addiction neuroscience, where laboratory findings often fail to connect to the spontaneous cognition and behavior that characterize the lived experience of substance use disorders [33, 34].

This study has several limitations. The stimulus was a single movie (Trainspotting), so generalization to other drug contexts and substances is untested. The sample was predominantly male (75% of the speech analyses, 69% of the brain-coupling analysis) and unequal in size (82 OUD, 34 HC), and nicotine use was almost exclusive to OUD, leaving sex differences, HC-specific topic effects, and the contribution of nicotine unresolved. Analyses were confined to a single dataset without independent training and testing samples, and the imaging subsample was further restricted to participants who completed both scanning sessions, for comparability with [14, 16], even though only session-1 data were analyzed here; replication in larger, multi-site cohorts with held-out validation is needed. Speech was collected about 45 minutes after viewing rather than immediately, which shows the effects survive a delay but adds variability from consolidation and from individual differences in recall. The clinical analyses were exploratory: correction was applied across regions within each measure rather than across the 13 measures, and the craving results therefore require independent confirmation. We did not test whether these linguistic markers predict treatment response or relapse, previously suggested for speech-based markers in cocaine addiction [15]. Finally, we analyzed linguistic content only, and the Speech Contrast is a single scalar chosen for interpretability that discards information present in the higher-dimensional sentence-level representations; acoustic and prosodic features, and explainability methods for the latent embedding dimensions, remain to be explored in future work.

Anchored to neural function and collected in minutes, the measure is a candidate for the low-burden outcome measures that addiction trials need [35], once it replicates in independent cohorts, and the same logic applies to any condition in which emotionally salient content is processed with a bias. Beyond addiction, a few minutes of unconstrained speech carries a trace of how the brain encoded an experience, and the route of that trace depends on what the experience meant to the speaker: in health it runs through frontoparietal cortex; in opioid addiction the prefrontal link weakens, the accumbens link reverses, and temporal cortex carries the trace instead.

## 4 Materials and Methods

### 4.1 Participants

Eighty-two individuals with OUD and 34 HC participants were recruited through a clinical trial evaluating intervention-related recovery outcomes in opioid addiction (NCT04112186), for which therapy-specific results will be reported separately. The Icahn School of Medicine at Mount Sinai Institutional Review Board approved all study procedures, which complied with all relevant ethical regulations, and all participants provided written informed consent. Participants completed all movie task procedures twice, during an initial baseline visit (Session 1) and approximately 15 weeks later (Session 2). During the interscan interval, OUD participants completed an 8-week group therapy program in addition to their regular inpatient treatment (see below). In this study, we utilized only data from session 1. For the analyses that involved fMRI data, we included only participants who completed the entire study, leaving a final sample of 35 OUD and 30 HC; this restriction keeps the sample identical to the fMRI cohort analyzed in [16] for comparability.

All OUD participants were recruited as inpatients enrolled in a treatment program that included prescription opioid medications for OUD (methadone: n=76, buprenorphine/naloxone: n=6) as well as regular therapy counseling and relapse prevention, trauma, and anger management courses, at a collaborating substance use rehabilitation organization in the New York City metropolitan area (Samaritan Daytop Village, Queens, NY). Age- and gender-matched HC participants were recruited through advertisements and word-of-mouth in the surrounding community.

Eligibility and participant characteristics were assessed during an initial screening with a comprehensive clinical interview administered by trained research coordinators supervised by clinical psychologists. Diagnostic evaluations included the Mini International Neuropsychiatric Interview, 7th Edition [36] and the Addiction Severity Index, 5th Edition [37]. All OUD participants met criteria for OUD with heroin as their primary substance of choice, and HC participants did not meet criteria for any current or past substance use disorder. Craving and withdrawal symptoms in OUD participants were assessed with the Heroin Craving Questionnaire (HCQ), a modified version of the Cocaine Craving Questionnaire [38], and the Short Opiate Withdrawal Scale (SOWS) [39], and their drug dependence severity was assessed with the Severity of Dependence Scale (SDS) [40]. Methadone or buprenorphine use was confirmed through urine toxicology at all screening and study visits. For all participants, current or past nicotine use and alcohol dependence were assessed with the Fagerström Test for Nicotine Dependence (FTND) [41] and the Short Michigan Alcohol Screening Test (SMAST) [42]. A brief medical examination including heart rate, blood pressure, alcohol concentration (with saliva test strips), breath carbon monoxide, and pregnancy testing when applicable (with urine test strips) was administered by trained research coordinators during each visit. Estimated verbal and nonverbal IQ were assessed with the reading subtest of the Wide Range Achievement Test-3 [43] and the Matrix Reasoning subtest of the Wechsler Abbreviated Scale of Intelligence [44]. Other neuropsychological measures included assessments for symptoms of depression (Beck’s Depression Inventory) [45] in the past two weeks and anxiety (Beck’s Anxiety Inventory) [46] in the past month. Group comparisons on demographic, neuropsychological, and substance use characteristics were performed using Mann-Whitney U tests for continuous variables and chi-squared tests for categorical variables in Python (scipy v1.15.2, statsmodels v0.14.4).

All participants were medically healthy at the time of study participation.

### 4.2 Experimental Protocol

During the session, participants viewed the first 17 minutes and 3 seconds of the movie Trainspotting with fMRI acquisition as described in our previous publication [14]. Scans were acquired on a Siemens 3T Skyra (Siemens, Erlangen, Germany) with a 32-channel head coil as part of a 2-hour MRI protocol that included other fMRI tasks [e.g., 47–49] and T1-, T2-, and diffusion-weighted scanning [e.g., 22, 50–52]. Details on the scanning procedures and MRI preprocessing steps can be found in our previous publication [14] and in the Supplementary Methods and Results.

Within 45 minutes of completing the MRI procedures, participants were instructed to recall the movie narrative and describe their experience of watching it while their speech was recorded. A research coordinator gave instructions as follows: “Please verbally recall the movie you just watched, in order, with as much detail as possible. The details are more important than the order. While recounting the movie, please also describe any thoughts, feelings, or emotions you may have experienced. Please tell us what these thoughts were and indicate the specific items in each of the scenes that made you have these feelings, and how strong your emotions were.” Participants performed the recall task uninterrupted, and no minimum or maximum time limit was imposed. Afterwards, participants completed a quality assurance questionnaire to assess their memory and comprehension of the movie, indicate whether they had seen it before, and rate their attention and understanding of the movie and the audio and visual quality as reported in King et al. [16].

Craving was measured at three points, as reported in King et al. [16]. Immediately before and immediately after the movie, participants rated their current heroin craving on a 0–9 scale (“Please rate how strong your desire for heroin is on a scale of 0–9”), giving the pre-scan and post-scan measures and their pre-to-post change. Following the speech task, participants rated craving on the same 0–9 scale for specific moments of the movie, sampled as 35 three-second clips taken at regular 30-second intervals across the segment; the mean and the maximum of these ratings are the scene-induced craving measures analyzed here (see *Drug-scene craving* below for how the drug-scene-restricted variants were derived). Because the scene ratings followed the recall task, they could not have shaped the speech analyzed here.

### 4.3 Speech Processing

Recordings were transcribed to text as in our previous publication, using OpenAI’s Whisper automated speech recognition model [53]. Transcribed texts underwent manual quality assurance processing for accuracy, including removal of interviewer speech and irrelevant participant speech (e.g., asking for clarification on the task instructions) [16]. The transcripts were split into sentences using NLTK’s sentence tokenizer (a Python package), then embedded into 384-dimensional vectors using *all-MiniLM-L6-v2* pretrained model from the SentenceTransformers framework for Bidirectional Encoder Representations from Transformers (BERT) sentence encoding [54]. Notably, this publicly available model runs on a standard computer with very low resources, making the method usable across clinical settings.

### 4.4 Topic Modeling and Producing Topic Labels

We applied BERTopic, a transformer-based topic modeling framework, to the full corpus of recall sentences pooled across all participants (3378 sentences), to identify recurring themes in participants’ recall narratives. BERTopic uses sentence embeddings in combination with a clustering algorithm and class-based TF–IDF (c-TF–IDF) to generate coherent topics from short text segments. Specifically, we first reduced the dimensionality of embeddings using Uniform Manifold Approximation and Projection. Clustering was performed using Hierarchical Density-Based Spatial Clustering of Applications with Noise at BERTopic’s default settings, allowing the model to flexibly identify dense semantic clusters while excluding noisy sentences. The full parameters used can be found in the Supplementary Methods. We trained our clustering model on all sentences from both groups together. This pipeline yielded a set of semantically coherent topics without requiring predefined categories, thereby reducing researcher bias.

For each sentence, the model produced soft topic membership scores across the identified topics. These scores were then aggregated by computing the mean score for each topic across all sentences from a participant to obtain individual-level representations. The resulting participant-level scores quantify the relative expression of each topic in a participant’s speech rather than representing true probabilities. Statistical comparisons of topic prevalence between groups were conducted using Welch’s unequal-variance *t*-statistic, pre-specified given the substantial heterogeneity of topic-prevalence variances between groups (OUD/HC variance ratios spanned 0.03 to 319 across topics), with a permutation test (10,000 label shuffles) to derive nominal p-values, followed by Benjamini– Hochberg false discovery rate (FDR) correction across all identified 21 topics; effect sizes are reported as Hedges’ g with bias-corrected-and-accelerated (BCa) bootstrap 95% confidence intervals (denoted *CI*_*BCa*_ throughout; 10,000 resamples). The confidence interval and p-value are computed by independent resampling procedures, so an interval that crosses zero and a p-value below .05 are not mutually exclusive for effects near the significance boundary. Potential confounding effects of demographic variables (Table 1) were examined via correlation analyses, and when significant relationships were detected, results were re-tested using Analysis of Covariance.

To facilitate interpretability, topic labels were assigned by **two human expert annotators** and **automated labeling** using the Gemini-1.5-pro-preview-0514 large language model. Representative sentences, those that were clustered for a certain topic (had highest membership score for this topic), from each topic were provided to the model with a standardized prompt (Supplementary Methods) to generate concise and informative labels. For topics containing sensitive drug-related content, automated labeling was restricted due to content filters, and labels were assigned exclusively by human experts. Human and model labels are reported side by side (Supplementary Table S1); because the human labels were assigned by consensus rather than independently, no inter-rater agreement statistic is reported.

### 4.5 Scene Similarity Analysis

#### 4.5.1 Movie Stimuli

The movie clip was annotated with precise temporal boundaries and detailed textual descriptions for each of 15 scenes. Scene content was classified as drug or non-drug following the criterion and blind-rating procedure of Kronberg et al. [14]: content was labeled **drug** if it contained any drugs, drug use, or a character who was high, and **nondrug** otherwise, with raters blind to the analyses. This process yielded 7 drug scenes (501 TRs) and 8 non-drug scenes (512 TRs), providing approximately balanced exposure across conditions.

#### 4.5.2 Sentence-to-Scene Mapping

Recall was analyzed at the sentence level, as an individual narrative typically references multiple scenes and sentence-level mapping permits attribution of each utterance to its corresponding scene. The movie was represented at whole-scene granularity because the analysis is defined over the 15 pre-specified scenes and each scene’s description serves as a single reliable target against which sentences are matched. Scene descriptions were embedded with the same model as the sentences (see Speech Processing), and each sentence was assigned to the scene with maximal **cosine similarity**:

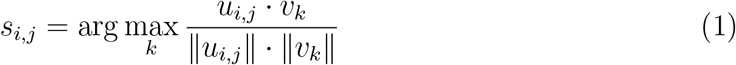

where *u*_*i,j*_ and *v*_*k*_ are the embeddings of sentence *j* from participant *i* and scene *k*, respectively (Fig. 4).

**Figure 4:**
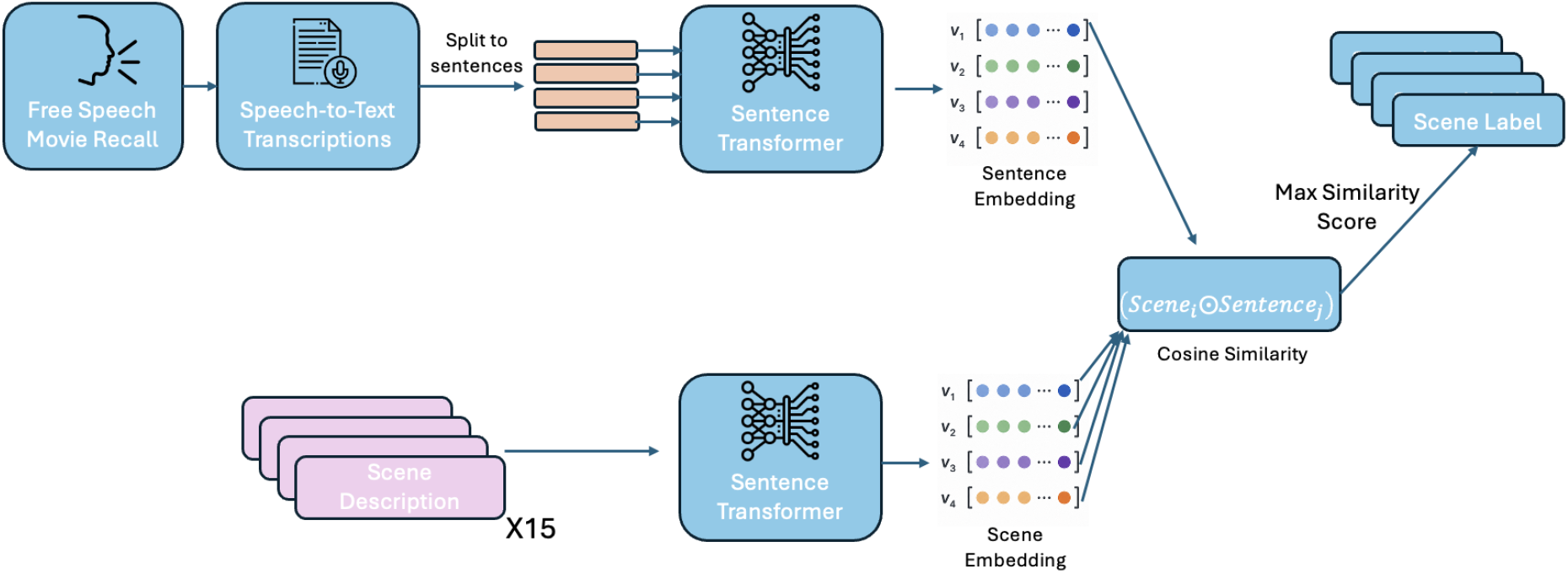
The scene similarity analysis. Free recall speech was transcribed and split into sentences, each encoded as a sentence embedding using a sentence transformer model. Scene descriptions were embedded using the same model, and each sentence was assigned to the scene with the highest cosine similarity score.

#### 4.5.3 Speech Contrast

For participant *i* with *n*_*i*_ sentences, the proportion of speech assigned to scene *j* was:

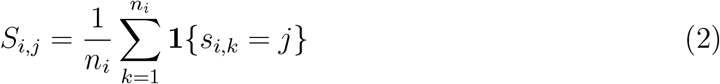

Proportions for each scene category were computed as:

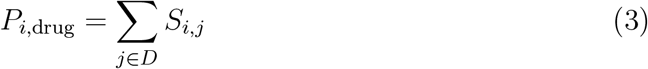

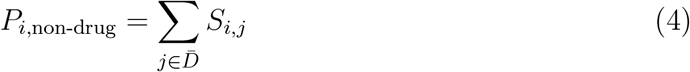

The **Speech Contrast** was then defined as:

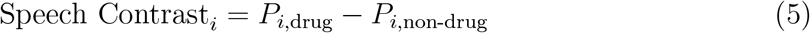

This participant-specific metric controls for variability in recall across scenes, ensuring balanced representation within each category. This full-sample scene-similarity analysis was repeated in the subset of participants with usable neuroimaging data (35 OUD, 30 HC; see Participants), as a robustness check against the smaller cohort used for the brain-speech coupling analysis below. Statistical comparisons of scene and drug/non-drug category proportions between groups were conducted using Welch’s t-statistic with a permutation test (10,000 label shuffles) to derive nominal p-values, followed by Benjamini– Hochberg FDR correction across the 15-scene family for individual-scene comparisons; effect sizes are reported as Hedges’ g with BCa bootstrap 95% confidence intervals (10,000 resamples). Full parameters are provided in the Supplementary Methods.

### 4.6 Brain-Speech Coupling Analysis

A distinctive feature of this design is that recall was collected after scanning rather than concurrently, linking speech and brain activity retrospectively: for each participant we related the Speech Contrast from post-scan recall to the Neural Contrast measured during movie viewing. This approach lets us test whether spontaneous recall reflects prior neural responsivity to drug-related content, without requiring participants to speak in the scanner.

#### 4.6.1 Neural Activation

BOLD signals were extracted from two atlases: the **Schaefer 400-parcel cortical atlas** (17-network parcellation) [17, 55] and the **Harvard-Oxford subcortical atlas** (15 structures) [18–21]; both Harvard–Oxford atlases were provided by the Center for Morphometric Analysis (Massachusetts General Hospital). The median activation for scene *j* in ROI *k* was computed identically in both atlases:

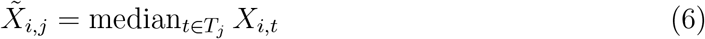

where *T*_*j*_ indexes all time points in scene *j*. Scene-category medians were then computed as:

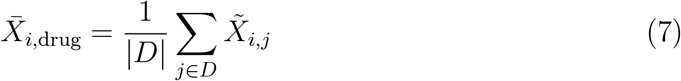

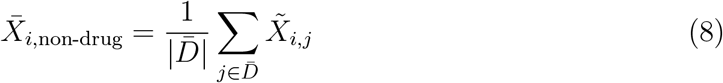

The **Neural Contrast** was defined as:

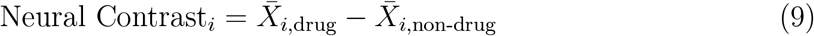

#### 4.6.2 Brain-Speech Coupling

Coupling between speech and neural activity was quantified by correlating the **Speech Contrast** with the **Neural Contrast** across participants using Pearson correlation, computed separately for the OUD and HC groups. Within-group significance was evaluated with a two-tailed permutation test (10,000 shuffles of the neural-contrast values, preserving the paired speech contrast). Between-group differences were quantified as a Fisher r-to-z difference and evaluated with a two-tailed permutation test (10,000 shuffles of group label across the pooled sample), since the direction of a between-group difference in coupling strength was not hypothesized a priori and could vary by region.

For the cortical atlas, multiple comparisons were controlled with **network-wise false discovery rate (FDR) correction** (Benjamini–Hochberg), applied separately within each of the Schaefer atlas’s 17 networks in each hemisphere (34 families) [55], to respect functional organization while retaining sensitivity in a modest sample. For the subcortical atlas, FDR correction was applied globally across all 15 structures. This approach balances statistical control with sensitivity in modest sample sizes (OUD: N = 35; HC: N = 30).

This analysis identifies brain regions where individual differences in the verbal elaboration of drug-related content are associated with differential neural responsivity to drug versus non-drug stimuli.

Because the group differences in the left nucleus accumbens and the left superior frontal gyrus ran in opposite directions, we tested that reversal directly. The interaction statistic is the difference between the two regions’ group differences in coupling, *D* = [*z*(*r*_OUD_) − *z*(*r*_HC_)]_accumbens_ − [*z*(*r*_OUD_) − *z*(*r*_HC_)]_SFG_, where *z* is the Fisher transform of the within-group correlation and each bracketed difference is divided by its standard error, the square root of 1*/*(*n*_OUD_ − 3) + 1*/*(*n*_HC_ − 3). *D* is therefore the difference between the two regions’ group-difference *z* statistics, not between the raw Fisher differences. Significance was assessed with a two-tailed group-label permutation test (10,000 shuffles), shuffling group across the pooled participants with group sizes preserved and each participant’s Speech Contrast and two Neural Contrast values kept together. Because both region-wise differences are computed in the same participants they are not independent, so an asymptotic test on their difference would be invalid; the permutation respects that dependence by construction. A stratified participant bootstrap (10,000 resamples) provides a confidence interval. As a parametric check, the data were stacked with two rows per participant and the Speech Contrast regressed on Neural Contrast × Group × Region by ordinary least squares with standard errors clustered by participant, the three-way term being the interaction of interest.

#### 4.6.3 Craving and drug-use measures

Within the OUD group we tested whether the 13 available drug-use, craving, and withdrawal measures (age of onset of regular use, duration of regular use, current abstinence, past 30-day use, HCQ, SOWS, SDS, craving rated before the scan, craving rated after the scan, the change in craving across the scan, and the mean, the maximum, and the session-to-session change of craving induced by the movie scenes) related to the Speech Contrast, to the Neural Contrast, or to brain–speech coupling in the five regions carrying the main results (left nucleus accumbens, LH_DefaultA_PFCd_3, LH_ Limbic_TempPole_7, LH_TempPar_1 and LH_TempPar_2). Moderation of coupling was tested as the interaction between the Neural Contrast and the measure, with all predictors standardized; the model was fitted in both directions, predicting the Neural Contrast and predicting the Speech Contrast, and supplemented by a median-split comparison of the within-group correlations, which is invariant to scale. Associations with the Speech Contrast and with the Neural Contrast alone were tested by rank correlation, in the full sample (*n* = 82) and in the imaging subsample (*n* = 35) where applicable. All tests were two-tailed permutation tests (10,000 shuffles), with Benjamini–Hochberg FDR correction across the five regions within each measure. Correction was not applied across the 13 measures, so these analyses are exploratory; every test is reported, including null results, in the Supplementary Results.

#### 4.6.4 Drug-scene craving

The scene-induced craving measure samples the movie on a fixed schedule: 35 three-second clips at regular 30-s intervals, each rated for whether it elicited craving and, if so, for intensity on a 0–9 scale. Because that schedule is independent of where the drug scenes fall, we recomputed the measure a priori over the drug scenes alone. An unendorsed clip was scored as 0. Clip *k* (*k* = 0 … 34) was mapped to movie TRs {30*k*, 30*k* + 1, 30*k* + 2}, and a clip was counted as a drug clip if any of its three TRs fell in the same majority-vote drug-TR set that defines the Neural and Speech Contrasts; 19 of the 35 clips qualified. The classification is stable under the clip-onset assumption: shifting every window by −3 TRs reclassifies two clips and by +3 TRs reclassifies none.

We derived six variables within the OUD group at session 1: the mean, the maximum, and the number of craving-endorsed drug clips; the SD and the range (maximum minus minimum) of intensity across all 35 clips; and the drug-clip mean minus the non-drug-clip mean, a within-subject drug-specific craving contrast. The range is not independent of the maximum: because an unendorsed clip scores 0, the minimum is 0 for nearly every participant, so the range reduces to the across-clip maximum (range vs. the original scene-craving maximum, *r* = 0.972 in the full sample and *r* = 0.998 in the imaging subsample). We therefore treat the range and the drug-clip maximum as one quantity throughout. Each was tested against the Speech Contrast (full sample and imaging subsample), the Neural Contrast, and brain–speech coupling (moderation, with a median-split sensitivity analysis) in the same five regions, using two-tailed permutation tests (10,000 shuffles) and rank correlations for the correlation tests. Following the a priori directional hypothesis motivating the analysis, no multiple-comparison correction was applied; every test performed is reported in the Supplementary Results and Supplementary Table S9. Per-clip ratings were unavailable for eight OUD participants, leaving *n* = 74 in the full sample and *n* = 32 in the imaging subsample.

## Data Availability

The de-identified participant-level Speech Contrast and ROI Neural Contrast values underlying each figure and table will be deposited in a public repository with a DOI upon journal publication. The recall transcripts, sentence embeddings and participant-level neuroimaging data were collected under clinical trial NCT04112186. They are available under controlled access through a data use agreement, on request to the authors, consistent with the trial consent and institutional review board approval. Controlled access is required because recall by inpatients about their own drug use, memories and treatment is potentially identifiable even with names removed, and sentence embeddings can be inverted to near-verbatim text. The raw audio recordings will not be shared. All analysis code will be made publicly available on GitHub upon journal publication.

## Data availability

The de-identified participant-level Speech Contrast and ROI Neural Contrast values underlying each figure and table will be deposited in a public repository with a DOI on publication. The recall transcripts, sentence embeddings, and participant-level neuroimaging data are available under controlled access through a data use agreement, consistent with the trial consent (NCT04112186) and institutional review board approval: recall by inpatients about their own drug use, memories, and treatment is potentially identifiable even with names removed, and sentence embeddings can be inverted to near-verbatim text. The raw audio recordings will not be shared.

## Code availability

All analysis code will be made publicly available on GitHub on publication and archived with a DOI together with the data above.

## Funding

This work was supported by the National Center for Complementary and Integrative Health grant R01AT010627 to R.Z.G.

## Author contributions

R.Z.G. designed the parent clinical study and the movie-recall paradigm and supervised data collection. S.P., R.T. and R.R. conceived the language-analysis approach. R.Z.G. formulated the iRISA-based hypotheses and the drug versus non-drug contrast and proposed the craving, interaction and robustness analyses. S.G.K. collected and curated the speech and neuroimaging data, including transcription quality assurance and preprocessing, and advised on data interpretation and statistics. S.P. developed the analysis code, performed the analyses, prepared the figures and wrote the first draft. S.P., R.Z.G. and R.R. wrote the manuscript; S.G.K. contributed text, R.T. contributed to the methodology, and N.C. advised on the writing; all three revised it. R.Z.G. acquired funding for the clinical study; R.R. acquired funding and computing resources for the language analyses. R.Z.G. and R.R. jointly supervised the project. All authors approved the final manuscript.

## Competing interests

The authors declare no competing interests.

## Notes

### Competing Interest Statement

The authors have declared no competing interest.

### Clinical Trial

NCT04112186

### Author Declarations

The Institutional Review Board of the Icahn School of Medicine at Mount Sinai gave ethical approval for this work.

