## Supplementary Material for "The path from brain to words is rerouted in opioid addiction"

### Supplementary Methods

**Topic modeling.** BERTopic (v0.17.0) was fit once, on the pooled corpus of 3,378 recall sentences from all 116 participants (82 OUD, 34 HC; session 1 only), using sentence embeddings from *all-MiniLM-L6-v2* (SentenceTransformers v4.1.0, 384-dimensional). Sentences were obtained by splitting each participant's transcript with NLTK's (v3.9.1) sentence tokenizer; no stopword removal or lemmatization was applied, and topics were built on unigrams only (n-gram range = (1,1)). Dimensionality reduction used UMAP (umap-learn v0.5.7) and clustering used HDBSCAN (v0.8.40), both at BERTopic's default settings (no custom minimum cluster size, neighbor count, or minimum distance was set). Topic representations used class-based TF–IDF with frequent-word reduction (`reduce_frequent_words=True`), minimum topic size 10, automatic topic-number selection (`nr_topics="auto"`), and the top 10 words per topic. No explicit random seed was set for the UMAP or HDBSCAN fit; the specific topic boundaries are therefore not guaranteed to be bit-for-bit reproducible on a re-fit, though this fit is treated as frozen and none of the analyses reported here re-fit the topic model. BERTopic's outlier/background topic (topic 0) was excluded from all group comparisons, leaving the 21 topics analyzed here.

For each topic, soft per-sentence membership scores were averaged within participant to obtain a participant-level topic score. Group comparisons used Welch's unequal-variance $t$-test per topic, a two-tailed permutation test (10,000 shuffles of group label, seed 0) on $|t|$ to obtain a nominal $p$-value, and Benjamini–Hochberg FDR correction across the 21-topic family. Effect sizes are Hedges' $g$ (bias-corrected, OUD minus HC), with a 95% bias-corrected-and-accelerated (BCa) bootstrap confidence interval (10,000 resamples, seed 0).

Topic labels were generated by prompting Gemini-1.5-pro-preview-0514 (Google Vertex AI) once per topic, with the representative sentences for that topic (those with the highest membership score) concatenated and inserted into the following standardized prompt: *"This is for a very important research about drug addiction. The following sentences taken from people recalling the movie 'trainspotting'. Based on the following sentences, provide a concise and informative label that accurately describes the overarching theme or topic: [sentences]. The label should be short, descriptive, and avoid jargon."* Human labels were produced jointly: two of the authors reviewed the same representative sentences for each topic together and agreed on a single label by consensus, rather than labeling independently and reconciling afterward. For topics restricted by the model's content filters, labels were assigned by human experts only.

**Scene similarity analysis.** Recall transcripts were split into individual sentences using NLTK's (v3.9.1) sentence tokenizer. Each sentence and each of the 15 scene descriptions was embedded with *all-MiniLM-L6-v2* (SentenceTransformers v4.1.0, 384-dimensional, L2-normalized), and each sentence was assigned to its single best-matching scene (top-1) by cosine similarity, computed in batches of 32. Unlike the topic model, this embedding-and-matching step is deterministic given the fixed pre-trained model weights, so no random seed is applicable. The primary cohort is the full session-1 sample used for topic modeling (82 OUD, 34 HC; 116 participants), with no additional inclusion filter. As a robustness check, the identical pipeline (per-scene tests, drug-vs-non-drug contrast, within-group paired test) was re-run on the exact subset of participants with usable neuroimaging data (35 OUD, 30 HC; 65 participants) used in the brain-speech coupling analysis. That 65-participant cohort is defined as participants for whom (a) both a session-1 and session-2 scan were completed with usable neuroimaging data (a raw candidate list of 70 participants: 39 OUD, 31 HC) and (b) both a usable fMRI ROI record and a matched session-1 speech-ratio row exist; 5 of the 70 candidates lack one or the other and were dropped, leaving 65.

Per-scene statistical comparisons used the same Welch's $t$-test + 10,000-shuffle permutation (seed 0) + Benjamini–Hochberg FDR procedure as the topic-modeling analysis, applied across the 15-scene family. The drug-vs-non-drug group contrast used the same Welch + permutation test applied directly to each participant's Speech Contrast value. Effect sizes for per-scene and drug-vs-non-drug group comparisons are Hedges' $g$ (bias-corrected, OUD minus HC) with a 95% BCa bootstrap confidence interval (10,000 resamples, seed 0), as for the topic-modeling analysis. Within-group drug-vs-non-drug comparisons used a paired $t$-test with a two-tailed permutation test (10,000 resamples), with Cohen's $d_z$ as the paired effect size.

**Covariate screening and ANCOVA.** For every result with a reported confound analysis, follow-up used the same two-step procedure. First, each candidate covariate (age, education, verbal IQ [WRAT reading], nonverbal IQ [WASI Matrix Reasoning], depression symptoms [BDI], anxiety symptoms [BAI], alcohol dependence [SMAST], scan interval, gender, race; nicotine/cigarette use was excluded) was tested for a group difference (Mann-Whitney U for continuous variables, chi-squared for categorical variables). Second, for variables differing by group, we tested their partial correlation with the outcome after residualizing both variables on group via ordinary least squares (Pearson correlation if both residuals were normally distributed by D'Agostino-Pearson test, Spearman otherwise; point-biserial correlation for categorical covariates). A potential covariate qualified for ANCOVA follow-up only if both tests were significant ($p < .05$); results with no qualifying covariate are reported as surviving by default. The topic-modeling and scene-similarity confound analyses use this partial-correlation ANCOVA. The brain–speech coupling analyses instead use the unified covariate model described next.

**Unified covariate model (brain–speech coupling).** The brain analyses used a single covariate model, applied identically to every region tested. We entered every variable that differed between OUD and HC within the fMRI subsample (education, verbal IQ, nonverbal IQ, depression symptoms [BDI], anxiety symptoms [BAI], alcohol dependence [SMAST]; Table S2), plus nicotine dependence (FTND). Age, gender, race and scan interval did not differ in this subsample and were excluded. To adjust a result, we residualized Neural Contrast and Speech Contrast on the covariates within each group by ordinary least squares, then re-ran the headline permutation test on the residuals. The adjusted $p$ is therefore directly comparable to the unadjusted $p$ in every test, within-group and between-group alike. We adjusted every region significant at $p < .05$ uncorrected. Listwise deletion left $N = 34$ OUD and $N = 29$ HC. One caveat: FTND spans 0–8 in OUD but is zero for 29 of the 30 HC participants, so the two groups are adjusted for unequal information.

**Brain-speech coupling.** fMRI acquisition and preprocessing followed our previous publication [1]; this pipeline operates on pre-extracted, parcel-averaged BOLD timeseries (1,019 TRs per session) rather than raw or minimally preprocessed images. For each participant and ROI, the timeseries was $z$-scored (after discarding the first 2 TRs of each session) prior to scene-wise aggregation. Neural Contrast and Speech Contrast were correlated (Pearson) separately within each group; within-group significance used a two-tailed permutation test (10,000 shuffles, seed 0) and the between-group comparison used a two-tailed label-permutation test (10,000 shuffles, seed 0) on the Fisher $z$-difference, as described in Methods. Analyses used the drug-versus-non-drug contrast throughout; this is the only contrast reported in the paper. Mean framewise displacement was not available for this dataset and was not included in the covariate set. Software: nibabel v5.3.2, nilearn v0.11.1, scipy v1.15.2, statsmodels v0.14.4 (Python 3.12.9).

### Supplementary Results

**Confound analyses (topic modeling).** To assess whether demographic differences could account for the topic effects, we screened each of the four FDR-surviving topics against the seven demographic/clinical variables that differed by group (race, education, verbal and nonverbal IQ, depression and anxiety symptoms, and alcohol dependence; Table 1) for a qualifying covariate: one that both differed by group and partially correlated with the topic controlling for group (see Methods). Only alcohol dependence qualified, correlating negatively with Topic 2 prevalence (partial $r_s = -0.25$, $p = .007$), opposite in sign to the OUD > HC group difference. An ANCOVA adjusting for alcohol dependence (ordinary least squares, unlike the permutation test behind the headline result) confirmed the Topic 2 group difference survived ($p = .003$); Topics 7, 11, and 13 had no qualifying covariates, so their headline results (Table 2) stand unadjusted.

**Confound analyses (scene similarity).** Using the same screening procedure, the full-sample drug-vs-non-drug category contrast (Speech Contrast) had one qualifying covariate, anxiety symptoms (BAI); an ANCOVA adjusting for it (ordinary least squares, unlike the permutation test behind the main-text result) confirmed the group difference survived ($p = .0159$; $N = 116$). Neither the fMRI-subset category contrast ($N = 65$) nor Scene 4 and Scene 8 (the only two individually significant scenes at uncorrected $p < .05$,

Table S1: Full BERTopic Topic Labeling and Group Comparisons (All 21 Topics)

| Topic # | Human Label | LLM Label | HC | OUD | $g$ [95% $CI_{BCa}$] | p (uncorr) | p (FDR) |
|---|---|---|---|---|---|---|---|
| 1 | Mother's Drug Consumption next to her Baby scene | Impact of Drug Addiction on Parental Care and Child Neglect | .024 | .017 | -0.15 [-0.68, 0.22] | .586 | .875 |
| **2** | **Personal Feeling and Identification with the movie** | **Emotional Identification and Reflection Through Shared Experiences of Addiction** | **.020** | **.035** | **0.47 [0.22, 0.67]** | **.003** | **.015** |
| 3 | Soccer Game Scene | N/A | .011 | .009 | -0.07 [-0.58, 0.27] | .747 | .923 |
| 4 | Complains about noise interfering with hearing | Impact of MRI Noise on Movie Comprehension | .006 | .009 | 0.30 [0.05, 0.47] | .050 | .151 |
| 5 | Movie Location - Scotland | Setting and Nationality: Perceptions of "Trainspotting" | .010 | .005 | -0.59 [-1.03, -0.16] | .078 | .194 |
| 6 | N/A - saying "I'm done" and the end as instructed | N/A | .004 | .013 | 0.34 [0.15, 0.50] | .032 | .112 |
| **7** | **Triggered "getting high" experiences and cravings** | **Memories and Emotional Responses to Drug Use in Film** | **.005** | **.011** | **0.45 [0.27, 0.58]** | **.001** | **.015** |
| 8 | "I can't remember" | Drug-Induced Memory Impairment | .009 | .010 | 0.09 [-0.40, 0.37] | .636 | .875 |
| 9 | "That's all i remember" | Fragmented Memories | .007 | .010 | 0.17 [-0.22, 0.39] | .308 | .588 |
| 10 | " I don't know" | Inability to Articulate Movie's Impact | .002 | .007 | 0.29 [0.16, 0.39] | .029 | .112 |
| **11** | **Personal usage craving and memories triggered by the movie** | **The Impact of "Trainspotting" on Drug Addiction Recovery: Triggers and Reminders** | **.005** | **.012** | **0.38 [0.25, 0.49]** | **<.001** | **.008** |
| 12 | Recalling the movie name | Movie Recall: Trainspotting | .004 | .012 | 0.22 [-0.10, 0.35] | .092 | .194 |
| **13** | **Desire for rehabilitation** | **Relapse and Resistance to Drug Addiction** | **.005** | **.009** | **0.50 [0.30, 0.66]** | **.003** | **.015** |
| 14 | A character describing the "high" feeling | N/A | .010 | .009 | -0.05 [-0.58, 0.25] | .801 | .932 |
| 15 | Expressing disgust (no context) | Negative Perception of Drug Use | .007 | .007 | -0.05 [-0.58, 0.39] | .848 | .932 |
| 16 | Expressing shock (no context) | Visceral Impact and Relatability of Drug Use Portrayal | .008 | .007 | -0.16 [-0.70, 0.34] | .666 | .875 |
| 17 | Describing unclear behavior in the movie | Memory Distortion of Drug Use in Film | .011 | .008 | -0.22 [-0.72, 0.24] | .561 | .875 |
| 18 | Third person perspective of enjoyment | Contradictory Perceptions of Pleasure and Addiction | .012 | .007 | -0.55 [-0.89, -0.13] | .090 | .194 |
| 19 | recalling a room or a house in the movie | Dilapidated Settings Reflecting Addiction | .009 | .010 | 0.03 [-0.41, 0.29] | .887 | .932 |
| 20 | N/A - saying "I'm done" and the end as instructed | Resolutions to Quit | .010 | .004 | -0.22 [-0.74, 0.22] | .568 | .875 |
| 21 | N/A - saying "I'm done" and the end as instructed | N/A - couldn't determine | .009 | .009 | -0.02 [-0.51, 0.31] | .939 | .939 |

**Note:** This table presents the comparison between human and LLM topic labeling along with their respective statistical analyses for all 21 topics identified by BERTopic. HC and OUD columns show group-mean topic-membership scores. $g$ is Hedges' g (OUD minus HC) with a BCa bootstrap 95% confidence interval (10,000 resamples); p (uncorr) is the nominal permutation-test p-value and p (FDR) is the Benjamini–Hochberg-corrected q-value across all 21 topics (see Methods). Bold rows indicate the four topics surviving FDR correction (p (FDR) < .05), presented in the main text (Table 2); topics 6 and 10 are nominally significant uncorrected but do not survive correction and are not bolded.

Table S2: Baseline demographic, neuropsychological, and drug use/addiction measures of participants in the final neuroimaging subsample (usable fMRI data, both scanning sessions completed)

| | HC<br>N = 30 | Opioid use disorder<br>OUD, N = 35 | Test statistic | *p* value |
|---|---|---|---|---|
| **Demographics** | | | | |
| Age, mean (SD) years | 40.0 (11.2) | 41.1 (10.7) | $U = 486$ | .612 |
| Gender, # men/women | 19/11 | 26/9 | $\chi^2 = 0.47$ | .494 |
| Race, # Black/White/multiracial or other | 8/14/8 | 7/20/8 | $\chi^2 = 0.75$ | .689 |
| Education, mean (SD) years | 16.2 (2.9) | 12.1 (2.6) | $U = 890$ | <.001 |
| **Neuropsychological tests** | | | | |
| WRAT - mean (SD) Reading standard score | 109.5 (6.6) | 96.1 (9.9) | $U = 908$ | <.001 |
| WASI - mean (SD) Matrix Reasoning scaled score | 11.7 (2.6) | 10.2 (2.9) | $U = 689$ | .030 |
| Depression symptoms, mean (SD) BDI score | 4.8 (9.8) | 11.9 (10.2) | $U = 213$ | <.001 |
| Anxiety symptoms, mean (SD) BAI score | 3.4 (6.8) | 8.4 (8.2) | $U = 264$ | <.001 |
| **Drug use and addiction variables** | | | | |
| Regular nicotine use, # current/past/never | 1/4/25 | 33/2/0 | – | – |
| Alcohol dependence, mean (SD) SMAST score | 0.3 (0.9) | 1.7 (2.0) | $U = 180$ | <.001 |
| *Heroin use disorder* | | | | |
| Age of onset, mean (SD) years | – | 25.5 (6.9) | – | – |
| Regular use, mean (SD) years | – | 10.7 (8.1) | – | – |
| Current abstinence, mean (SD) days | – | 220.1 (231.5) | – | – |
| Past 30-day use, mean (SD) days | – | 0.2 (0.7) | – | – |
| Craving symptoms, mean (SD) HCQ score | – | 35.5 (13.7) | – | – |
| Withdrawal symptoms, mean (SD) SOWS score | – | 2.6 (3.0) | – | – |
| Severity of dependence, mean (SD) SDS score | – | 10.5 (4.1) | – | – |

**Notes**: The table presents mean and standard deviation (SD) or quantities for baseline sample characteristics. Test statistics were derived from a Mann-Whitney U test ($U$) for continuous variables and chi-squared test ($\chi^2$) for categorical variables. Since only one HC participant reported current nicotine use and only 2 OUD participants reported non-current nicotine use, no statistical test was performed on this measure.

Table S3: Brain regions with uncorrected brain–speech coupling in OUD participants (drug vs. non-drug contrast)

| **Network** | **ROI** | **r** | **p (uncorr)** | **p (FDR)** |
|---|---|---|---|---|
| Control A | LH_ContA_PFCl_2 | .42 | .014 | – |
| Control A | LH_ContA_IPS_5 | .37 | .032 | – |
| Default Mode B | LH_DefaultB_Temp_2 | .42 | .013 | – |
| Default Mode B | RH_DefaultB_AntTemp_1 | −.34 | .050 | – |
| Dorsal Attention A | LH_DorsAttnA_SPL_5 | .43 | .009 | – |
| Dorsal Attention A | LH_DorsAttnA_SPL_6 | .33 | .049 | – |
| Dorsal Attention B | RH_DorsAttnB_PostC_2 | −.36 | .036 | – |
| **Limbic** | **LH_Limbic_TempPole_7** | **.46** | **.006** | **.042** |
| Salience/VenAttn A | RH_SalVentAttnA_ParOper_3 | −.39 | .022 | – |
| Salience/VenAttn A | RH_SalVentAttnA_Ins_1 | −.44 | .012 | – |
| **Temporal Parietal** | **LH_TempPar_2** | **.44** | **.007** | **.035** |
| **Temporal Parietal** | **LH_TempPar_1** | **.41** | **.012** | **.035** |
| Temporal Parietal | RH_TempPar_1 | −.45 | .008 | – |

**Notes**: All 13 regions with a significant correlation ($p < .05$, uncorrected) between Speech Contrast and Neural Contrast in OUD participants are shown; the within-group test is two-tailed (see Methods), so both positive and negative correlations are tested (positive $r$: greater neural differentiation of drug scenes with greater drug-scene recall; negative $r$: the reverse). Bold rows indicate the three ROIs surviving network-wise FDR correction, presented in the main text; negative correlations were tested but none reached significance after FDR correction.

Table S4: Brain regions with significant FDR-corrected brain–speech coupling in HC participants (drug vs. non-drug contrast)

| **Network** | **ROI** | **x** | **y** | **z** | **r** | **p (FDR)** |
|---|---|---|---|---|---|---|
| Control A | LH_ContA_IPS_5 | −33 | −46 | 41 | .68 | .001 |
| | LH_ContA_IPS_4 | −45 | −41 | 47 | .61 | .003 |
| | LH_ContA_PFCl_1 | −48 | 35 | 10 | .47 | .039 |
| | LH_ContA_PFCl_4 | −44 | 20 | 27 | .45 | .045 |
| Control B | LH_ContB_PFClv_3 | −28 | 57 | 13 | .55 | .010 |
| | LH_ContB_IPL_3 | −42 | −52 | 49 | .54 | .010 |
| | LH_ContB_PFCd_1† | −30 | 14 | 57 | .46 | .028 |
| | LH_ContB_PFClv_2 | −28 | 58 | −1 | .45 | .028 |
| | LH_ContB_PFClv_1 | −42 | 49 | −7 | .44 | .028 |
| | LH_ContB_IPL_2 | −53 | −50 | 45 | .39 | .050 |
| Default Mode A | LH_DefaultA_PFCd_2 | −18 | 36 | 48 | .62 | .002 |
| | LH_DefaultA_PFCd_3 | −22 | 19 | 52 | .55 | .024 |
| Default Mode B | LH_DefaultB_PFCd_3 | −22 | 50 | 31 | .62 | .004 |
| | LH_DefaultB_PFCl_1 | −41 | 19 | 48 | .53 | .036 |

**Notes**: All 14 ROIs with significant Speech Contrast–Neural Contrast coupling in HC participants after network-wise FDR correction; all are in the left hemisphere, and all surviving correlations were positive. The within-group test is two-tailed (see Methods): negative correlations were tested but none reached significance after FDR correction in either group. † This ROI fell just short of significance under the unified covariate model (adjusted $p = .052$; see Supplementary Results). $x$, $y$, $z$ are the MNI coordinates of each parcel's centroid (centre of gravity of its voxels in the Schaefer-400 17-network atlas); because the analysis is parcel-wise, every voxel in a parcel carries the same value, so no peak voxel is defined.

Table S5: Brain regions with uncorrected brain–speech coupling in HC participants (drug vs. non-drug contrast)

| Network | ROI | x | y | z | r | p (uncorr) | p (FDR) |
|---|---|---|---|---|---|---|---|
| **Control A** | **LH_ContA_IPS_5** | **−33** | **−46** | **41** | **.68** | **<.001** | **.001** |
| **Control A** | **LH_ContA_IPS_4** | **−45** | **−41** | **47** | **.61** | **<.001** | **.003** |
| **Control A** | **LH_ContA_PFCl_1** | **−48** | **35** | **10** | **.47** | **.009** | **.039** |
| **Control A** | **LH_ContA_PFCl_4** | **−44** | **20** | **27** | **.45** | **.014** | **.045** |
| Control A | RH_ContA_IPS_4 | 36 | −45 | 45 | .39 | .032 | – |
| Control A | RH_ContA_PFCl_3 | 47 | 29 | 28 | .39 | .037 | – |
| **Control B** | **LH_ContB_PFClv_3** | **−28** | **57** | **13** | **.55** | **.002** | **.010** |
| **Control B** | **LH_ContB_IPL_3** | **−42** | **−52** | **49** | **.54** | **.002** | **.010** |
| **Control B** | **LH_ContB_PFCd_1**† | **−30** | **14** | **57** | **.46** | **.011** | **.028** |
| **Control B** | **LH_ContB_PFClv_2** | **−28** | **58** | **−1** | **.45** | **.014** | **.028** |
| **Control B** | **LH_ContB_PFClv_1** | **−42** | **49** | **−7** | **.44** | **.014** | **.028** |
| Control B | RH_ContB_PFCld_4 | 34 | 16 | 55 | .41 | .021 | – |
| **Control B** | **LH_ContB_IPL_2** | **−53** | **−50** | **45** | **.39** | **.030** | **.050** |
| Control B | RH_ContB_PFClv_3 | 42 | 51 | −6 | .38 | .032 | – |
| Control B | LH_ContB_PFCmp_1 | −4 | 28 | 47 | .38 | .036 | – |
| Control B | RH_ContB_PFCmp_1 | 5 | 28 | 48 | .37 | .043 | – |
| **Default Mode A** | **LH_DefaultA_PFCd_2** | **−18** | **36** | **48** | **.62** | **<.001** | **.002** |
| **Default Mode A** | **LH_DefaultA_PFCd_3** | **−22** | **19** | **52** | **.55** | **.003** | **.024** |
| Default Mode A | LH_DefaultA_PFCd_1 | −25 | 28 | 43 | .48 | .008 | – |
| **Default Mode B** | **LH_DefaultB_PFCd_3** | **−22** | **50** | **31** | **.62** | **<.001** | **.004** |
| **Default Mode B** | **LH_DefaultB_PFCl_1** | **−41** | **19** | **48** | **.53** | **.003** | **.036** |
| Default Mode B | LH_DefaultB_PFCd_4 | −8 | 43 | 51 | .44 | .014 | – |
| Default Mode B | LH_DefaultB_PFCv_4 | −48 | 28 | 0 | .44 | .016 | – |
| Default Mode B | LH_DefaultB_Temp_5 | −61 | −35 | −3 | .44 | .016 | – |
| Default Mode B | LH_DefaultB_Temp_1 | −43 | 13 | −33 | .40 | .027 | – |
| Default Mode B | LH_DefaultB_PFCv_5 | −53 | 19 | 11 | .37 | .045 | – |
| Default Mode B | LH_DefaultB_PFCd_1 | −4 | 51 | 28 | .37 | .049 | – |
| Default Mode C | RH_DefaultC_IPL_1 | 48 | −64 | 22 | −.43 | .017 | – |
| Dorsal Attention A | LH_DorsAttnA_SPL_6 | −36 | −52 | 56 | .50 | .005 | – |
| Dorsal Attention A | RH_DorsAttnA_SPL_5 | 31 | −64 | 53 | .48 | .008 | – |
| Dorsal Attention A | RH_DorsAttnA_SPL_7 | 34 | −51 | 54 | .40 | .032 | – |
| Dorsal Attention B | LH_DorsAttnB_FEF_2 | −26 | 0 | 55 | .40 | .026 | – |
| Salience/VenAttn A | LH_SalVentAttnA_Ins_2 | −40 | −14 | −2 | −.38 | .038 | – |
| Salience/VenAttn A | RH_SalVentAttnA_ParMed_1 | 8 | 2 | 43 | −.41 | .024 | – |
| Salience/VenAttn B | RH_SalVentAttnB_PFCmp_1 | 7 | 35 | 25 | .36 | .045 | – |
| Somatomotor B | LH_SomMotB_Aud_6 | −59 | −37 | 16 | −.36 | .048 | – |
| Somatomotor B | RH_SomMotB_S2_10 | 59 | 1 | 10 | −.38 | .039 | – |

**Notes**: All 37 regions with a significant correlation ($p < .05$, uncorrected) between Speech Contrast and Neural Contrast in HC participants are shown; the within-group test is two-tailed (see Methods), so both positive and negative correlations are tested (positive $r$: greater neural differentiation of drug scenes with greater drug-scene recall; negative $r$: the reverse). Bold rows indicate the 14 ROIs surviving network-wise FDR correction, presented in the main text; all are in the left hemisphere; negative correlations were tested but none reached significance after FDR correction. † This ROI fell just short of significance under the unified covariate model (adjusted $p = .052$; see Supplementary Results) and is retained here for completeness. $x$, $y$, $z$ are the MNI coordinates of each parcel's centroid (centre of gravity of its voxels in the Schaefer-400 17-network atlas); because the analysis is parcel-wise, every voxel in a parcel carries the same value, so no peak voxel is defined.

Table S6: Brain regions with uncorrected group differences in brain–speech coupling (HC vs. OUD, drug vs. non-drug contrast)

| **Network** | **ROI** | **x** | **y** | **z** | **Fisher $z$** | **p (uncorr)** | **p (FDR)** | **Direction** |
|---|---|---|---|---|---|---|---|---|
| Control A | LH_ContA_IPS_4 | −45 | −41 | 47 | −2.44 | .026 | – | HC > OUD |
| Control B | RH_ContB_PFCld_1 | 39 | 33 | 38 | −1.99 | .038 | – | HC > OUD |
| Control B | LH_ContB_PFCd_1 | −30 | 14 | 57 | −1.74 | .007 | – | HC > OUD |
| **Default Mode A** | **LH_DefaultA_PFCd_3** | **−22** | **19** | **52** | **−3.07** | **.001** | **.022** | **HC > OUD** |
| Default Mode A | LH_DefaultA_PFCd_2 | −18 | 36 | 48 | −2.62 | .006 | – | HC > OUD |
| Default Mode A | LH_DefaultA_PFCd_1 | −25 | 28 | 43 | −2.29 | .026 | – | HC > OUD |
| Default Mode B | RH_DefaultB_PFCd_3 | 5 | 45 | 40 | −2.59 | .013 | – | HC > OUD |
| Default Mode B | LH_DefaultB_PFCd_3 | −22 | 50 | 31 | −2.34 | .031 | – | HC > OUD |
| Default Mode B | LH_DefaultB_PFCd_4 | −8 | 43 | 51 | −2.09 | .029 | – | HC > OUD |
| Default Mode C | LH_DefaultC_PHC_1 | −21 | −21 | −26 | +1.49 | .048 | – | OUD > HC |
| Dorsal Attention A | RH_DorsAttnA_SPL_5 | 31 | −64 | 53 | −2.98 | .006 | – | HC > OUD |
| Dorsal Attention A | RH_DorsAttnA_SPL_7 | 34 | −51 | 54 | −2.18 | .013 | – | HC > OUD |
| Dorsal Attention A | LH_DorsAttnA_SPL_5 | −29 | −58 | 51 | +1.46 | .040 | – | OUD > HC |
| Somatomotor B | LH_SomMotB_Aud_6 | −59 | −37 | 16 | +2.45 | .007 | – | OUD > HC |
| Somatomotor B | LH_SomMotB_Aud_11 | −61 | −17 | 19 | +1.69 | .030 | – | OUD > HC |
| Visual Central | RH_VisCent_ExStr_4 | 19 | −88 | −13 | −1.67 | .045 | – | HC > OUD |

**Notes**: All 16 ROIs with a significant difference in Speech Contrast–Neural Contrast coupling between groups ($p < .05$, uncorrected) are shown. Negative $z$ values indicate stronger coupling in the HC group; positive $z$ values indicate stronger coupling in the OUD group. The bold row indicates the one ROI surviving network-wise FDR correction, presented in the main text. $x$, $y$, $z$ are the MNI coordinates of each parcel's centroid (centre of gravity of its voxels in the Schaefer-400 17-network atlas); because the analysis is parcel-wise, every voxel in a parcel carries the same value, so no peak voxel is defined.

Table S7: Subcortical brain–speech coupling, all 15 structures (drug vs. non-drug contrast, Harvard–Oxford atlas)

| | **OUD** | | | **HC** | | | **Group difference** | | |
|---|---|---|---|---|---|---|---|---|---|
| **Structure** | **r** | **p** | **p (FDR)** | **r** | **p** | **p (FDR)** | **z** | **p** | **p (FDR)** |
| **Left Accumbens** | **0.34** | **.048** | **.361** | **−0.52** | **.004** | **.054** | **+3.55** | **<.001** | **.001** |
| Right Thalamus | 0.06 | .734 | .926 | −0.27 | .139 | .347 | +1.30 | .145 | .363 |
| Right Hippocampus | −0.04 | .799 | .926 | −0.34 | .066 | .329 | +1.19 | .127 | .363 |
| Left Hippocampus | 0.11 | .515 | .917 | −0.29 | .122 | .347 | +1.57 | .105 | .363 |
| Left Pallidum | −0.23 | .189 | .777 | 0.30 | .102 | .347 | −2.08 | .054 | .363 |
| Right Amygdala | 0.04 | .803 | .926 | −0.44 | .018 | .132 | +1.95 | .080 | .363 |
| Left Thalamus | −0.15 | .393 | .917 | 0.09 | .660 | .982 | −0.89 | .331 | .709 |
| Left Caudate | 0.35 | .040 | .361 | −0.00 | .999 | .999 | +1.40 | .388 | .727 |
| Right Pallidum | 0.10 | .550 | .917 | −0.09 | .653 | .982 | +0.73 | .519 | .865 |
| Right Putamen | −0.00 | .978 | .978 | −0.10 | .608 | .982 | +0.36 | .717 | .913 |
| Brainstem | 0.13 | .469 | .917 | 0.04 | .842 | .982 | +0.35 | .702 | .913 |
| Right Caudate | 0.22 | .207 | .777 | 0.06 | .742 | .982 | +0.61 | .730 | .913 |
| Left Amygdala | −0.08 | .648 | .926 | −0.02 | .916 | .982 | −0.23 | .806 | .930 |
| Right Accumbens | −0.03 | .882 | .945 | −0.04 | .856 | .982 | +0.04 | .962 | .962 |
| Left Putamen | −0.13 | .448 | .917 | −0.10 | .612 | .982 | −0.14 | .919 | .962 |

**Notes**: All 15 subcortical structures tested are shown. All p-values are two-tailed permutation p-values, Benjamini–Hochberg FDR-corrected across the 15 structures (see Methods). The bold row indicates the one structure surviving correction on the group difference, presented in the main text.

Table S8: Drug-use, craving, and withdrawal measures against the Speech Contrast, the Neural Contrast, and brain–speech coupling in the OUD group (all 13 measures)

**(a) Speech Contrast**

| | Full sample | | | Imaging subsample | | |
|---|---|---|---|---|---|---|
| **Measure** | $\rho$ | **p** | **n** | $\rho$ | **p** | **n** |
| Age of onset | 0.02 | .870 | 82 | **0.38** | **.027** | 35 |
| Duration of use | −0.08 | .487 | 79 | **−0.38** | **.028** | 34 |
| Current abstinence | −0.12 | .293 | 82 | 0.05 | .770 | 35 |
| Past 30-day use | −0.11 | .336 | 82 | −0.10 | .575 | 35 |
| HCQ | 0.19 | .093 | 81 | 0.06 | .718 | 35 |
| SOWS | 0.17 | .135 | 81 | 0.19 | .275 | 35 |
| SDS | 0.05 | .646 | 82 | −0.04 | .811 | 35 |
| Craving, pre-scan | 0.15 | .187 | 80 | 0.13 | .463 | 34 |
| Craving, post-scan | 0.07 | .548 | 80 | 0.07 | .669 | 34 |
| Craving, pre-to-post change | −0.15 | .180 | 80 | −0.11 | .510 | 34 |
| Scene craving, mean | 0.19 | .080 | 81 | −0.07 | .707 | 35 |
| Scene craving, maximum | 0.22 | .053 | 82 | 0.15 | .378 | 35 |
| Scene craving, session change | 0.04 | .826 | 40 | 0.16 | .360 | 35 |

**(b) Brain–speech coupling: moderation fitted in both model directions (strongest of the five main-result regions)**

| | | Neural Contrast as outcome | | Speech Contrast as outcome | |
|---|---|---|---|---|---|
| **Measure** | **Region** | $\beta$ | **q** | $\beta$ | **q** |
| Age of onset | L Acc | −0.27 | .388 | −0.47 | .0505 |
| Duration of use | L Acc | 0.05 | .838 | −0.12 | .457 |
| Current abstinence | L Acc | −0.15 | .622 | −0.26 | .732 |
| Past 30-day use | TempPar_2 | −0.87 | .506 | −0.30 | .647 |
| HCQ | PFCd_3 | −0.31 | .343 | −0.21 | .965 |
| SOWS | L Acc | 0.03 | .816 | −0.08 | .827 |
| SDS | TempPole_7 | 0.24 | .203 | 0.16 | .550 |
| Craving, pre-scan | TempPole_7 | 0.30 | .045‡ | 0.10 | .984 |
| Craving, post-scan | TempPole_7 | 0.26 | .142 | 0.11 | .761 |
| Craving, pre-to-post change | L Acc | 0.07 | .914 | −0.12 | .977 |
| Scene craving, mean | L Acc | −0.05 | .709 | −0.03 | .861 |
| Scene craving, maximum | TempPole_7 | 0.22 | .927 | 0.27 | .700 |
| Scene craving, session change | L Acc | 0.15 | .366 | 0.06 | .920 |

**(c) Neural Contrast and median-split coupling (strongest of the five main-result regions)**

| | Neural Contrast | | | Coupling (median split) | |
|---|---|---|---|---|---|
| **Measure** | **Region** | **r** | **q** | **Region** | **q** |
| Age of onset | L Acc | 0.16 | .506 | L Acc | .053 |
| Duration of use | L Acc | −0.32 | .113 | L Acc | .696 |
| Current abstinence | TempPar_2 | 0.44 | .052 | PFCd_3 | .107 |
| Past 30-day use | TempPar_2 | −0.37 | .145 | L Acc | .902 |
| HCQ | L Acc | 0.13 | .744 | L Acc | .894 |
| SOWS | L Acc | 0.24 | .607 | L Acc | .938 |
| SDS | PFCd_3 | 0.22 | .381 | L Acc | .603 |
| Craving, pre-scan | TempPole_7 | −0.29 | .453 | TempPole_7 | .830 |
| Craving, post-scan | TempPole_7 | −0.31 | .392 | L Acc | .733 |
| Craving, pre-to-post change | PFCd_3 | 0.26 | .696 | L Acc | .966 |
| Scene craving, mean | L Acc | 0.14 | .648 | L Acc | .587 |
| Scene craving, maximum | L Acc | 0.20 | .906 | L Acc | .937 |
| Scene craving, session change | TempPole_7 | 0.36 | .160 | TempPole_7 | .063 |

**Notes**: All 13 drug-use, craving, and withdrawal measures available in the OUD group are shown; all tests are within OUD at session 1. Panel (a) reports rank correlations with the Speech Contrast; p-values are two-tailed permutation p-values (10,000 shuffles), uncorrected, as the Speech Contrast is a single outcome. Panels (b) and (c) report, for each measure, the strongest of the five main-result regions in each block; $q$ is the Benjamini–Hochberg FDR-corrected permutation p-value across those five regions within that measure (see Methods). Correction was not applied across the 13 measures, so these analyses are exploratory. Bold entries in panel (a) are significant at the uncorrected level. Panel (b) reports the moderation fitted in both directions at the same region, so the two fits can be compared cell by cell; no $q$ reaches .05 in the speech-outcome fit, the smallest being age of onset at the accumbens ($q = .0505$, shown to four decimals for that reason). The median-split block in panel (c) compares within-group correlations and is therefore invariant to model direction. ‡ Craving before the scan reached $q = .045$ in two regions in the neural-outcome fit, LH_Limbic_TempPole_7 ($\beta = 0.30$; listed) and LH_TempPar_2 ($\beta = 0.40$), but in neither the speech-outcome fit nor the median-split test; it is not interpreted as a coupling effect. Region abbreviations: L Acc, left nucleus accumbens; PFCd_3, LH_DefaultA_PFCd_3 (left superior frontal gyrus); TempPole_7, LH_Limbic_TempPole_7; TempPar_1 and TempPar_2, LH_TempPar_1 and LH_TempPar_2. Age of onset and duration refer to regular use; the scene-craving measures are the fixed-schedule movie ratings described in Methods. Sample sizes vary by measure with item-level missingness; the session-change measure of scene-induced craving was available in 40 participants in the full sample. HCQ, Heroin Craving Questionnaire; SOWS, Short Opiate Withdrawal Scale; SDS, Severity of Dependence Scale.

Table S9: Drug-scene craving (a priori, uncorrected): six drug-clip-restricted craving variables against the Speech Contrast, the Neural Contrast, and brain–speech coupling in the OUD group

**(a) Speech Contrast**

| | Full sample | | | Imaging subsample | | |
|---|---|---|---|---|---|---|
| **Variable** | $\rho$ | **p** | **n** | $\rho$ | **p** | **n** |
| Drug-clip maximum† | **0.29** | **.011** | 74 | 0.17 | .352 | 32 |
| Range across clips† | **0.28** | **.016** | 74 | 0.14 | .439 | 32 |
| Drug − non-drug mean | **0.27** | **.023** | 74 | 0.20 | .263 | 32 |
| SD across clips | 0.23 | .050 | 74 | 0.03 | .876 | 32 |
| Drug-clip mean | 0.21 | .078 | 74 | −0.05 | .802 | 32 |
| Drug clips endorsed (count) | 0.16 | .159 | 74 | −0.08 | .664 | 32 |

**(b) Neural Contrast and brain–speech coupling (strongest of the five main-result regions)**

| | Neural Contrast | | | Coupling (moderation) | | | Coupling (median split) | |
|---|---|---|---|---|---|---|---|---|
| **Variable** | **Region** | **r** | **p** | **Region** | $\beta$ | **p** | **Region** | **p** |
| Drug-clip maximum† | L Acc | 0.27 | .136 | TempPar_1 | −0.22 | .380 | TempPar_1 | .158 |
| Range across clips† | L Acc | 0.23 | .190 | TempPar_2 | −0.21 | .406 | TempPar_1 | .158 |
| Drug − non-drug mean | TempPar_2 | 0.24 | .191 | TempPar_1 | −0.10 | .532 | TempPar_1 | .184 |
| SD across clips | L Acc | 0.18 | .334 | TempPar_2 | −0.13 | .529 | PFCd_3 | .565 |
| Drug-clip mean | TempPar_1 | 0.19 | .296 | TempPar_1 | −0.19 | .256 | TempPar_1 | .265 |
| Drug clips endorsed (count) | TempPole_7 | −0.17 | .346 | TempPar_2 | −0.31 | .137 | TempPar_1 | .265 |

**Notes**: Scene-induced craving recomputed over the 19 of 35 movie clips overlapping the drug scenes that define the Neural and Speech Contrasts (see Methods); all tests are within OUD at session 1. All p-values are two-tailed permutation p-values (10,000 shuffles). **No multiple-comparison correction is applied**: these tests were specified a priori on a directional hypothesis, and every test performed is reported here or in the Supplementary Results. Panel (b) reports, for each variable, the strongest of the five main-result regions in each block; no test in any of these three blocks reached $p < .05$, including all 30 moderation and all 30 median-split tests. Bold entries in panel (a) are significant at the uncorrected level. Sample sizes are smaller than elsewhere in the paper ($n = 74$ full sample, $n = 32$ imaging subsample) because per-clip ratings were unavailable for eight OUD participants. Range across clips is the maximum minus the minimum intensity over all 35 clips; drug − non-drug mean is the within-subject difference between the drug-clip and non-drug-clip means. † These two variables are one quantity, not two independent findings: an unendorsed clip scores 0, so the minimum is 0 for nearly every participant and the range reduces to the across-clip maximum (range vs. the original scene-craving maximum, $r = 0.972$ full sample, $r = 0.998$ imaging subsample; drug-clip maximum vs. the same original maximum, $r = 0.941$). Of the three significant variables, only the drug − non-drug contrast is genuinely distinct from the original scene-craving measures ($r = 0.700$ with the original mean, $r = 0.634$ with the original maximum). Region abbreviations as in Table S8.

in both the full sample and the fMRI subset) had a qualifying covariate, so all three headline results stand unadjusted. No individual scene reached the 15-scene FDR threshold in either cohort (main text).

**fMRI-subset replication (scene similarity).** The full-sample drug-vs-non-drug group contrast ($g = 0.68$, 95% $CI_{BCa}$ [0.28, 1.10]; OUD mean difference = 0.268, HC mean difference = 0.003; $t = -3.24$, permutation $p = .002$; $N = 116$) replicated in the smaller fMRI subset ($g = 0.65$, 95% $CI_{BCa}$ [0.08, 1.08]; OUD mean difference = 0.237, HC mean difference = −0.015; $t = -2.58$, permutation $p = .012$; $N = 65$). The within-group paired drug-vs-non-drug effect, significant only in OUD in the full sample (OUD permutation $p < .001$; HC permutation $p = .972$), was likewise OUD-specific in the fMRI subset (OUD permutation $p = .0003$; HC permutation $p = .846$).

**Confound analyses (brain-speech coupling).** We re-tested every FDR-significant ROI under the unified covariate model (see Methods), which applies the same seven covariates to every region and adjusts by residualizing within group before re-running the headline permutation test. Both between-group FDR-significant results survived: the left accumbens ($z = 3.55 \rightarrow 3.61$, adjusted permutation $p = .0001$) and the left superior frontal gyrus in Default Mode A (LH_DefaultA_PFCd_3; $z = -3.07 \rightarrow -2.53$, adjusted $p = .004$). All three OUD within-group FDR-significant ROIs survived: LH_TempPar_2 ($r = .44 \rightarrow .51$, adjusted $p = .001$), LH_Limbic_TempPole_7 ($r = .46 \rightarrow .52$, adjusted $p = .002$), and LH_TempPar_1 ($r = .41 \rightarrow .38$, adjusted $p = .025$). Of the 14 HC within-group FDR-significant ROIs, 13 survived; one, LH_ContB_PFCd_1, fell just short of significance after adjustment ($r = .46 \rightarrow .37$, adjusted $p = .052$; flagged with † in Supplementary Tables S4 and S5). Dropping FTND from the model left every verdict unchanged except this same borderline ROI, which then crossed the threshold (adjusted $p = .047$); no other FDR-significant result was affected. Full per-ROI results are provided as source data.

**Range-restriction check (brain-speech coupling).** To confirm that the weaker within-group coupling in OUD participants (and hence the HC > OUD group differences) did not arise from restricted variance in the OUD group, we compared group spreads and re-tested the differences under range-matching, focusing on the regions carrying the group-difference claim: the left superior frontal gyrus (Default Mode A; LH_DefaultA_PFCd_3) and the left nucleus accumbens. Because this check concerns the dispersion of the per-participant contrast values, it is independent of the tail and permutation choices used for the coupling significance tests. Speech Contrast spread was comparable between groups (OUD $SD = 0.341$, HC $SD = 0.433$; Fligner–Killeen $p = .34$, Levene $p = .29$), as was Neural Contrast spread in the cortical region (Fligner $p = .26$). In the left nucleus accumbens the OUD group was in fact *more* dispersed than HC ($SD = 0.119$ vs. 0.073; Levene $p = .009$, Fligner $p = .012$): the opposite of range restriction. Restricting the wider group to the narrower group's range preserved every group difference with its sign intact (all range-matched $p < .01$; the accumbens difference strengthened, Fisher $z = +3.55 \rightarrow +3.94$), and a group-label permutation test confirmed each difference (left superior frontal gyrus $p = .001$; left accumbens $p = .0001$). The coupling asymmetry therefore reflects a genuine group difference rather than compressed OUD variance.

**Craving and drug-use measures (brain–speech coupling).** All 13 drug-use, craving, and withdrawal measures were tested against the Speech Contrast (full sample, $n = 82$, and imaging subsample, $n = 35$), the Neural Contrast, and brain–speech coupling in the five main-result regions, with FDR correction across the five regions within each measure. Two interactions reached the corrected threshold, both moderations by craving rated before the scan, and both only with the Neural Contrast as the model outcome: LH_Limbic_TempPole_7 ($\beta = 0.30$, $q = .045$) and LH_TempPar_2 ($\beta = 0.40$, $q = .045$; $n = 34$). Because a moderation model's outcome is not an arbitrary choice, we fitted both directions. With the Speech Contrast as outcome neither effect was present ($\beta = 0.10$, $p = .61$; $\beta = 0.01$, $p = .96$), and a median-split comparison of the within-group correlations, which is invariant to scale and therefore arbitrates between the two fits, supported neither ($q = .83$, $q = .84$). Across the median split the correlation itself changed little while the spread of the Neural Contrast grew and that of the Speech Contrast shrank, so the slope difference is carried by the ratio of standard deviations rather than by coupling. The corresponding effect for craving rated after the scan did not survive correction in either direction ($q = .14$ and $q = .76$). We report these as null. Craving induced by the drug scenes did not moderate coupling in any region: the nucleus accumbens interaction was flat and nominally opposite in sign, and the superior frontal gyrus, though in the predicted direction, was far from significance ($p = .70$). The only nominal association of scene-induced craving anywhere was with the Speech Contrast in the full sample ($\rho = 0.215$, $p = .053$). No measure survived correction against accumbens or prefrontal coupling; the nearest trends were accumbens coupling in participants with earlier onset of use ($q = .053$) and control-like positive prefrontal coupling with longer abstinence ($q = .107$). Within the imaging subsample, later onset of regular use was associated with a higher Speech Contrast ($\rho = 0.38$, $p = .027$) and longer duration of use with a lower one ($\rho = -0.38$, $p = .028$); neither association was present in the full sample and neither survived correction. All remaining tests were null.

**Drug-scene craving (a priori, uncorrected).** Restricting the scene-craving measure to the 19 of 35 clips overlapping the drug scenes yielded six variables, each tested against four outcome blocks within the OUD group without multiple-comparison correction (see Methods); all tests performed are reported here and in Supplementary Table S9. Against the Speech Contrast in the full sample ($n = 74$), three of the six were significant: the drug-clip maximum ($\rho = 0.290$, $p = .011$), the across-clip range ($\rho = 0.277$, $p = .016$), and the drug-minus-non-drug mean ($\rho = 0.266$, $p = .023$); the across-clip SD was borderline ($\rho = 0.229$, $p = .050$), and the drug-clip mean ($\rho = 0.207$, $p = .078$) and endorsed-clip count ($\rho = 0.165$, $p = .159$) were null. In the imaging subsample ($n = 32$) all six were null ($|\rho| \leq 0.20$, $p \geq .26$; strongest, drug-minus-non-drug, $\rho = 0.201$, $p = .263$). No test in the remaining blocks reached $p < .05$ even uncorrected. Across the 30 Neural Contrast tests (six variables $\times$ five regions) the largest was the nucleus accumbens with the drug-clip maximum, in the predicted direction but not significant ($r = 0.265$, $p = .136$). Across the 30 moderation tests and 30 median-split tests, none reached $p < .05$; every accumbens moderation was far from significance ($p \geq .47$), and the accumbens median splits leaned in the predicted direction only trivially (drug-minus-non-drug: $r = 0.40$ in the high half vs. $r = 0.22$ in the low half, $p = .457$), while the superior frontal gyrus splits leaned against the prediction. Two of the three significant variables are not independent findings. Because an unendorsed clip scores 0, the minimum is 0 for nearly every participant and the across-clip range reduces to the across-clip maximum, correlating with the original scene-craving maximum at $r = 0.972$ in the full sample and $r = 0.998$ in the imaging subsample. The drug-clip maximum tracks that same original maximum nearly as closely (below), so the maximum and the range are effectively one variable. The one significant variable genuinely distinct from the original measures is the drug-minus-non-drug contrast, which overlaps them only moderately ($r = 0.700$ with the original mean, $r = 0.634$ with the original maximum) and is also the most interpretable, being a within-subject contrast. The restriction otherwise did not materially change the measure: because 19 of the 35 clips already overlapped the drug scenes, the drug-clip mean correlated with the original scene-craving mean at $r = 0.961$ (Spearman $\rho = 0.973$) and the drug-clip maximum with the original maximum at $r = 0.941$ (Spearman $\rho = 0.953$; $n = 74$). The standardized 30-s sampling therefore was not diluting drug-scene craving, and clip sampling does not explain the absent accumbens association.
